# Interdisciplinary Inquiry via PanelGPT: Application to Explore Chatbot Application in Sports Rehabilitation

**DOI:** 10.1101/2023.07.23.23292452

**Authors:** Joseph C. McBee, Daniel Y. Han, Li Liu, Leah Ma, Donald A. Adjeroh, Dong Xu, Gangqing Hu

## Abstract

**Background:** ChatGPT showcases exceptional conversational capabilities and extensive cross-disciplinary knowledge. In addition, it possesses the ability to perform multiple roles within a single chat session. This unique multi-role-playing feature positions ChatGPT as a promising tool to explore interdisciplinary subjects.

**Objective:** The study intended to guide ChatGPT for interdisciplinary exploration through simulated panel discussions. As a proof-of-concept, we employed this method to evaluate the advantages and challenges of using chatbots in sports rehabilitation.

**Methods:** We proposed a model termed PanelGPT to explore ChatGPTs’ knowledge graph on interdisciplinary topics through simulated panel discussions. Applied to “chatbots in sports rehabilitation”, ChatGPT role-played both the moderator and panelists, which included a physiotherapist, psychologist, nutritionist, AI expert, and an athlete. We act as the audience posed questions to the panel, with ChatGPT acting as both the panelists for responses and the moderator for hosting the discussion. We performed the simulation using the ChatGPT-4 model and evaluated the responses with existing literature and human expertise.

**Results:** Each simulation mimicked a real-life panel discussion: The moderator introduced the panel and posed opening/closing questions, to which all panelists responded. The experts engaged with each other to address inquiries from the audience, primarily from their respective fields of expertise. By tackling questions related to education, physiotherapy, physiology, nutrition, and ethical consideration, the discussion highlighted benefits such as 24/7 support, personalized advice, automated tracking, and reminders. It also emphasized the importance of user education and identified challenges such as limited interaction modes, inaccuracies in emotion-related advice, assurance on data privacy and security, transparency in data handling, and fairness in model training. The panelists reached a consensus that chatbots are designed to assist, not replace, human healthcare professionals in the rehabilitation process.

**Conclusions:** Compared to a typical conversation with ChatGPT, the multi-perspective approach of PanelGPT facilitates a comprehensive understanding of an interdisciplinary topic by integrating insights from experts with complementary knowledge. Beyond addressing the exemplified topic of chatbots in sports rehabilitation, the model can be adapted to tackle a wide array of interdisciplinary topics within educational, research, and healthcare settings.

## Introduction

The sports industry, a significant economic contributor in the U.S., is projected to generate $83.1 billion of revenue in 2023 ^1^. Concurrently, sports/recreation-related injuries are prevalent, with an estimated rate of 34 per 1000 individuals, which accumulates to an annual total of 8.6 million cases ^2^. Sports rehabilitation, aiming to facilitate full recovery, minimize sports downtime, and prevent future injuries, results from a coordinated effort between the athlete and healthcare professionals across various disciplines ^3^. However, the rehabilitation process often spans over a lengthy period and demands expensive medical and psychological support, making it inaccessible for many patients. In recent year, the integration of artificial intelligence (AI) in sports medicine have shown promise in enhancing both the accessibility to service and efficacy of treatment outcomes ^4^. However, the use of chatbots in assisting sports rehabilitation is still in its formative stages, with many of the potential benefits and pitfalls yet to be explored and understood.

ChatGPT, a sophisticated large language model (LLM)-based chatbot, is capable of human-like dialogues ^5^. Trained on a vast dataset spanning a wide range of disciplines, ChatGPT has an attractive feature: multi-role-playing, which allows the chatbot to assume the roles of several discipline-specific experts at the same time. This unique feature inspired us to propose a model for exploring interdisciplinary topics through a simulated panel discussion, where ChatGPT assumes the roles of a moderator and various experts on the panel. We employed this model to navigate the multifaceted interdisciplinary landscape of chatbot-assisted sports rehabilitation and summarized our findings.

## Methods

We proposed a model (named as PanelGPT) to explore interdisciplinary topics through a simulated panel discussion powered by ChatGPT (**Figure 1A**). In this model, ChatGPT assumes the roles of both the moderator and panel experts at varying times, while a human operator, representing people/humans in the audience, poses questions and sends reminders to the moderator or the panelists. Questions from the human operator are directly copied and pasted into the chat session, with ChatGPT determining which panel member(s) should respond. If the discussion stalls at any point, the human operator prompt the moderator or panelists to continue by sending reminders. After each round of discussion, the moderator summarizes the comments before moving to solicit the next question from the audience. Upon conclusion of the panel discussion, the chatbot’s responses and suggestions are summarized and evaluated through the literature or human expert opinions.

**Figure 1.**
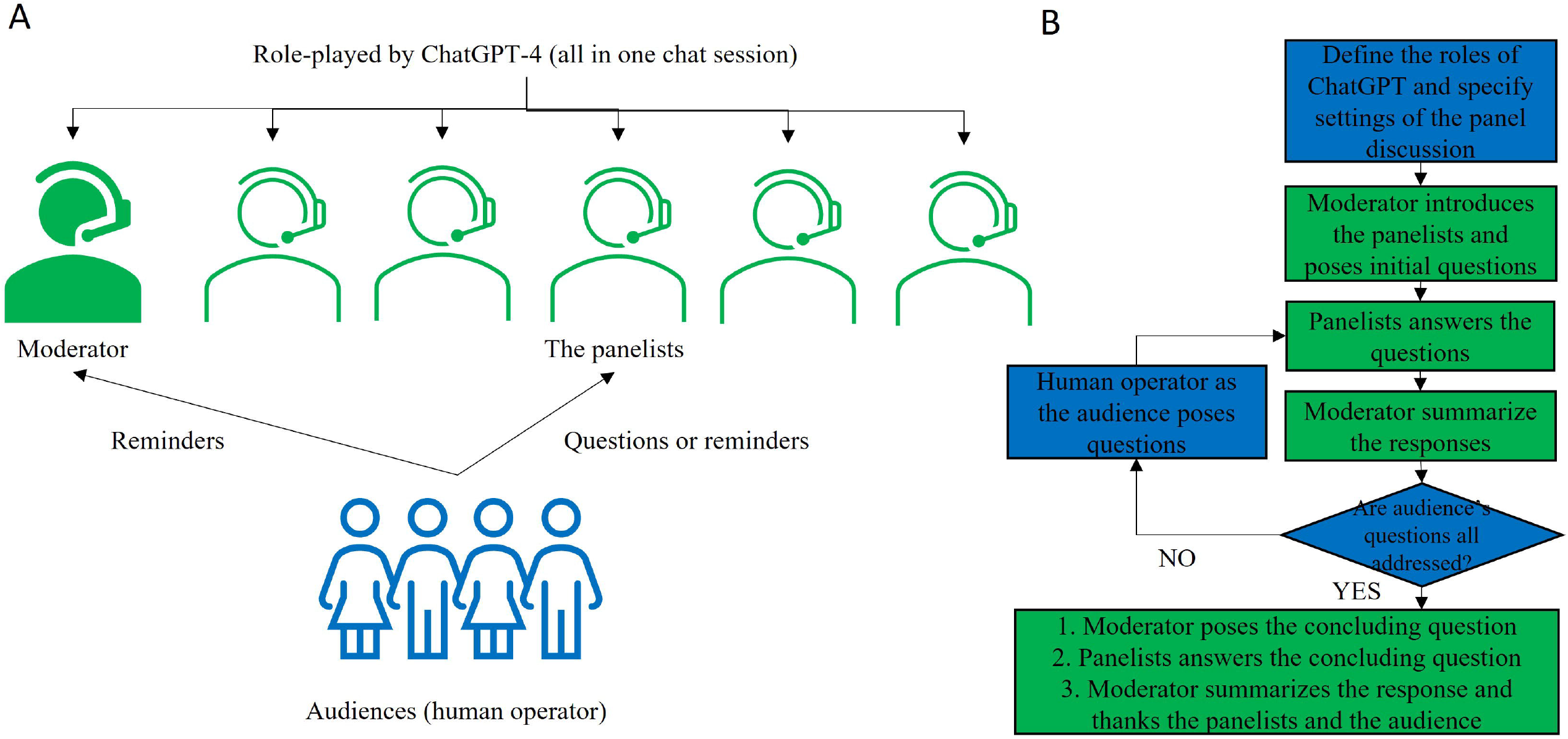
An overview of the PanelGPT model for a ChatGPT-simulated panel discussion (A), and a flow chart that delineates the process of the simulation (B).

As an illustrative example, we applied PanelGPT to explore the use of chatbots in sports rehabilitation. The simulated panel comprised four experts representing essential disciplines related to the topic: a physiotherapist, a psychologist, a nutritionist, and an AI expert specializing in clinical applications. In addition, a virtual athlete who had successfully recovered from a severe injury participated in the panel. Initially, we formulated four main questions based on personal experience and literature review. After reviewing the responses from pilot simulations, we added two more questions (**Table 1**). During one of the pilot simulations, ChatGPT autonomously introduced opening questions, which we subsequently included in the final simulations. This finding also prompted us to instruct the chatbot to ask concluding questions at the end of each simulation.

**Table 1.**
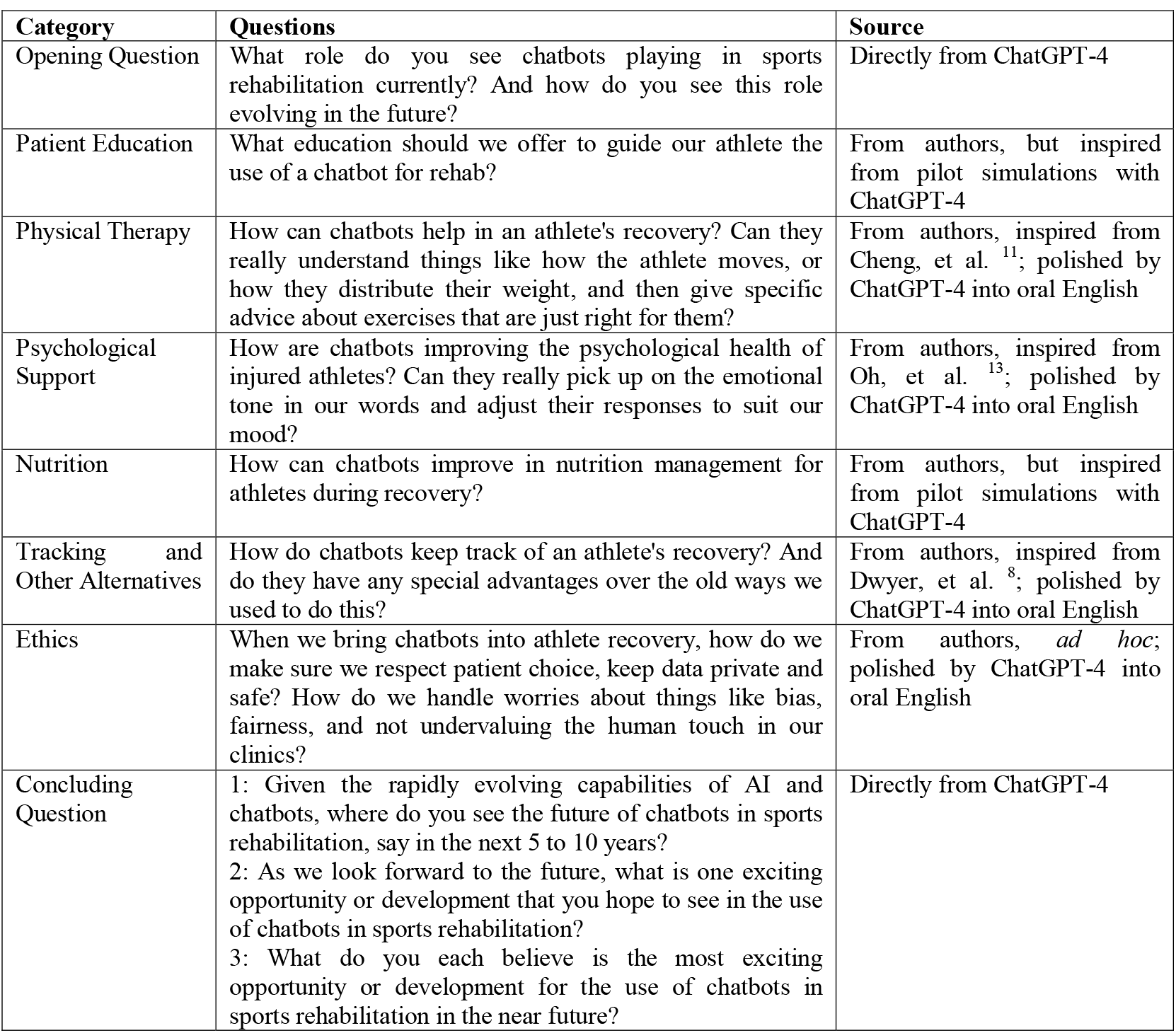
Questions designed for the simulated panel discussion on “chatbots in sports rehabilitation”.

| Category | Questions | Source |
| --- | --- | --- |
| Opening Question | What role do you see chatbots playing in sports rehabilitation currently? And how do you see this role evolving in the future? | Directly from ChatGPT-4 |
| Patient Education | What education should we offer to guide our athlete the use of a chatbot for rehab? | From authors, but inspired from pilot simulations with ChatGPT-4 |
| Physical Therapy | How can chatbots help in an athlete's recovery? Can they really understand things like how the athlete moves, or how they distribute their weight, and then give specific advice about exercises that are just right for them? | From authors, inspired from Cheng, et al. <sup>11</sup>; polished by ChatGPT-4 into oral English |
| Psychological Support | How are chatbots improving the psychological health of injured athletes? Can they really pick up on the emotional tone in our words and adjust their responses to suit our mood? | From authors, inspired from Oh, et al. <sup>13</sup>; polished by ChatGPT-4 into oral English |
| Nutrition | How can chatbots improve in nutrition management for athletes during recovery? | From authors, but inspired from pilot simulations with ChatGPT-4 |
| Tracking and Other Alternatives | How do chatbots keep track of an athlete's recovery? And do they have any special advantages over the old ways we used to do this? | From authors, inspired from Dwyer, et al. <sup>8</sup>; polished by ChatGPT-4 into oral English |
| Ethics | When we bring chatbots into athlete recovery, how do we make sure we respect patient choice, keep data private and safe? How do we handle worries about things like bias, fairness, and not undervaluing the human touch in our clinics? | From authors, *ad hoc*; polished by ChatGPT-4 into oral English |
| Concluding Question | 1: Given the rapidly evolving capabilities of AI and chatbots, where do you see the future of chatbots in sports rehabilitation, say in the next 5 to 10 years?<br>2: As we look forward to the future, what is one exciting opportunity or development that you hope to see in the use of chatbots in sports rehabilitation?<br>3: What do you each believe is the most exciting opportunity or development for the use of chatbots in sports rehabilitation in the near future? | Directly from ChatGPT-4 |

To avoid misunderstanding, our focus is not on the use of ChatGPT to provide sports rehabilitation advice. Instead, we centered on the use of ChatGPT to drive a panel discussion entitled “chatbots in sports rehabilitation”. The prompts used to steer the final simulations are detailed in **Table 2**. A flow chart that outlines the process of the simulation is shown in **Figure 1B**. Initially, we instructed ChatGPT to undertake multiple roles and specified other settings in the simulation (**Table 2**). The moderator was initially prompted to introduce the panelists and kickoff the discussion with opening questions. Following the responses to these initial questions from the panelists, the moderator was then tasked to summarize the responses and then open the platform for questions from the audience. In response to this, the human operator copied each audience question directly into the chat session, allowing ChatGPT to autonomously select which expert should respond. After each round of questions and answers, the moderator was prompted to summarize the responses and then call for the next question. This process iterated until all audience questions had been addressed. Finally, at the conclusion of the panel discussion, the moderator was asked to propose a closing question and provide a summary of the responses. Additional prompts were introduced as needed to ensure the smooth progression of the panel discussion (**Table 2**). We repeated the simulation three times using ChatGPT-4 (May 24 version).

**Table 2.**
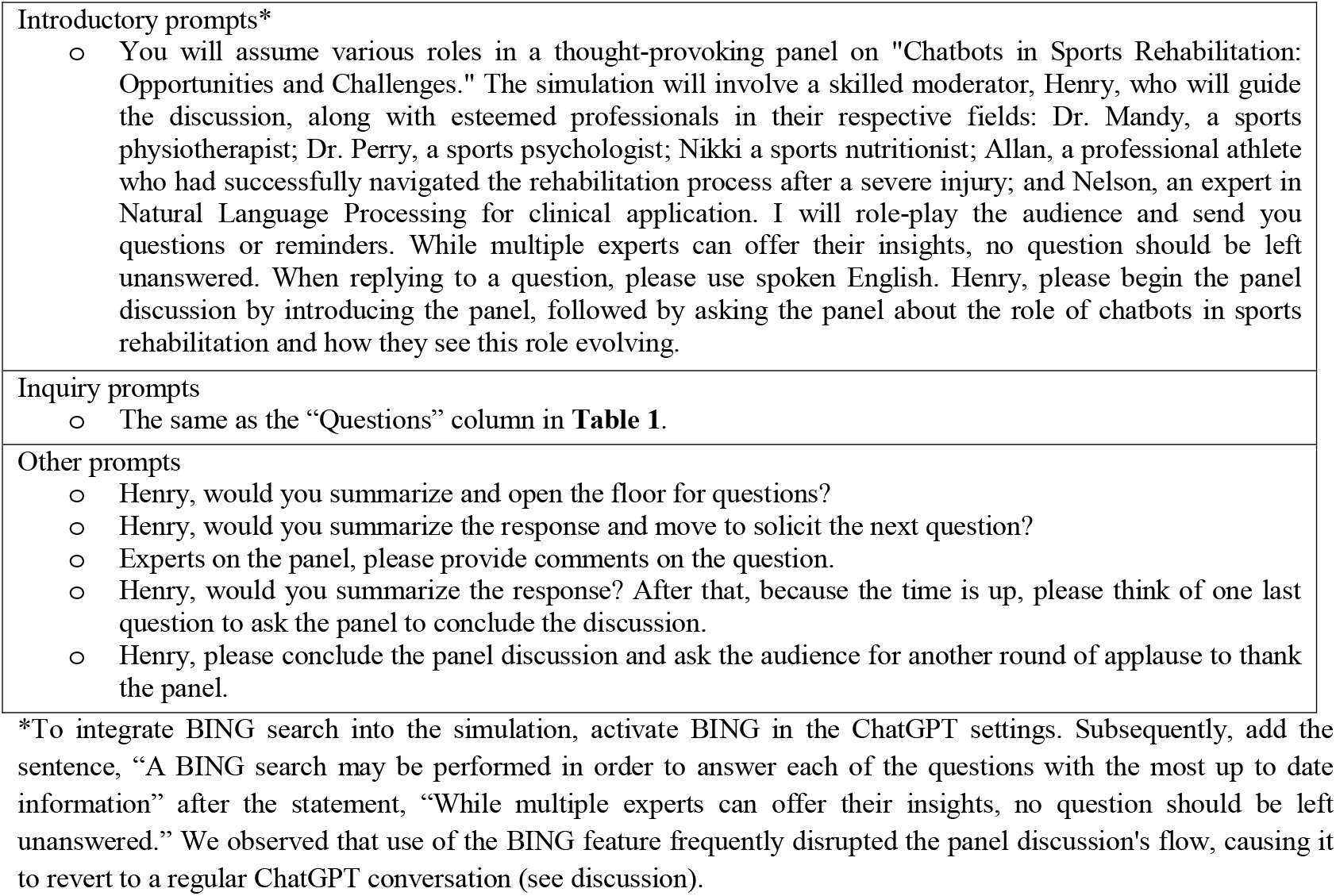
Prompts used to steer the simulated panel discussion.

## Results

The prompts used in the simulated panel discussion and the corresponding scripts are accessible in **Supplementary Files 1 to 3** (audio version available upon request). As expected, two or more experts responded to each question. The experts generally offered insights from the standpoint of their respective fields of expertise (**Table 3**). We evaluated the responses and added references when necessary for clarification. Major findings were compiled and summarized below.

**Table 3.**
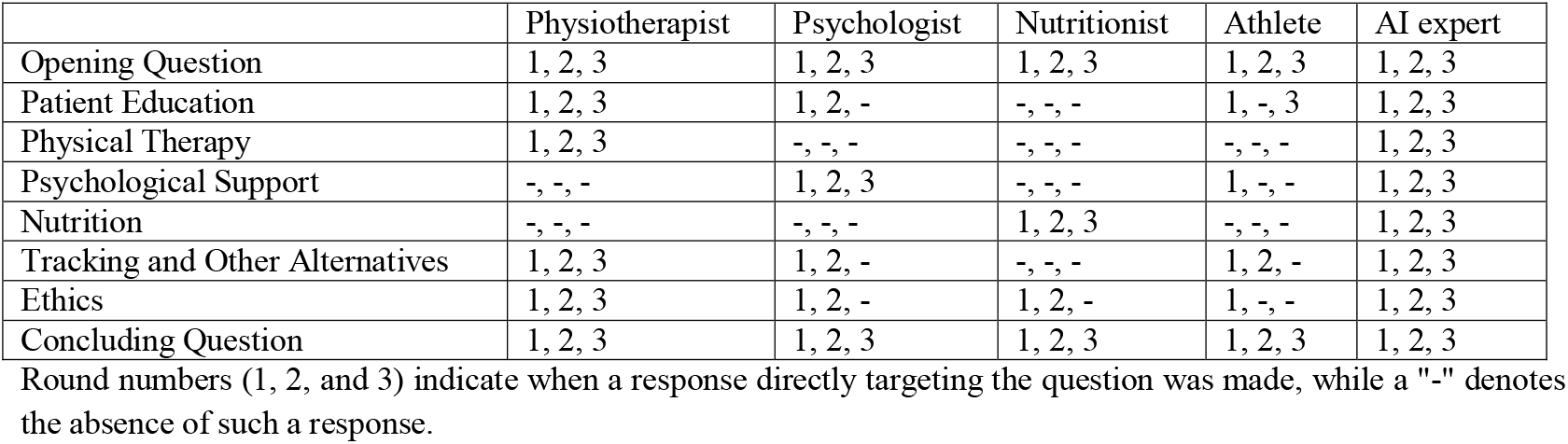
Records on direct responses to questions during the simulation.

|  | Physiotherapist | Psychologist | Nutritionist | Athlete | AI expert |
| --- | --- | --- | --- | --- | --- |
| Opening Question | 1, 2, 3 | 1, 2, 3 | 1, 2, 3 | 1, 2, 3 | 1, 2, 3 |
| Patient Education | 1, 2, 3 | 1, 2, - | -, -, - | 1, -, 3 | 1, 2, 3 |
| Physical Therapy | 1, 2, 3 | -, -, - | -, -, - | -, -, - | 1, 2, 3 |
| Psychological Support | -, -, - | 1, 2, 3 | -, -, - | 1, -, - | 1, 2, 3 |
| Nutrition | -, -, - | -, -, - | 1, 2, 3 | -, -, - | 1, 2, 3 |
| Tracking and Other Alternatives | 1, 2, 3 | 1, 2, - | -, -, - | 1, 2, - | 1, 2, 3 |
| Ethics | 1, 2, 3 | 1, 2, - | 1, 2, - | 1, -, - | 1, 2, 3 |
| Concluding Question | 1, 2, 3 | 1, 2, 3 | 1, 2, 3 | 1, 2, 3 | 1, 2, 3 |

Round numbers (1, 2, and 3) indicate when a response directly targeting the question was made, while a "-" denotes the absence of such a response.

### Opening Question

The simulated panel discussion began with introductions and requests for the panelists’ perspectives on the role of chatbots in sports rehabilitation, to which all panel members provided responses (**Table 3**). The ensuing dialogue identified chatbots as round-the-clock support systems, adept at monitoring, offering reminders, consulting, and nurturing a positive mindset in athletes during their recovery. Similar observations are reported for orthopedic patients ^6-8^. Looking into the future and consistent with expectations, it was suggested that chatbots might grow increasingly adept at analyzing biomechanical data, emotional indicators, and nutritional needs, thus providing personalized feedback that helps athletes better comprehend their bodies and healing journeys.

### Patient Education

The conversation pinpointed several key factors in educating athletes on the use of chatbots for rehabilitation. Both the athlete and the psychologist touched the importance of understanding the benefits of using a chatbot, such as serving as a readily available source for advice and mental support ^9^. The AI expert emphasized on educating transparency: how data is collected, processed, stored, and protected. Effective communication with a Chatbot is a non-trivial task ^10^. The physiotherapist focused on guiding users on how to effectively interact with the chatbot for the most useful responses, and how to interpret the responses. The discussion also underscored that the chatbot system is designed to enhance recovery, not to replace human touch. Through education, athletes need to be able to identify situations that call for direct communication with healthcare professionals.

### Physical Therapy

The primary focus of the questions was on the chatbot’s potential in facilitating physical therapy by analyzing movements and weight distributions ^11^. Relevant responses were from the physiotherapist and the AI expert, who both acknowledged that current chatbots primarily interact with users through text and voice, which restricts their direct applicability to the question. Yet, the AI expert envisioned the possibility of integrating chatbots, wearables, cameras, and smart devices to analyze an athlete’s movement patterns and providing real-time, personalized feedback. A good example as we noticed in the literature is computer vision-based analysis that has been applied to monitor and improve sports performance ^12^. The AI expert further highlighted that the accuracy of this application depends on the size and quality of the training data, as well as advances in AI technologies like machine learning and computer vision.

### Psychological Support

This round of discussion explored the role of chatbots in analyzing emotional cues via sentiment analysis, a technique previously shown to enhance patient satisfaction in several medical chatbot applications ^13-15^ and other applications ^16^. The panel’s responses aligned with the existing literature: by delivering tailored responses to emotions, the chatbots offer athletes with emotional support and reduce their feelings of being isolated. Nevertheless, the panel did not explore the impact of chatbots on psychological outcome measures, such as improvements in communication skills, cognitive level, motivation, and abilities in coping with the injury. Both the psychologist and the AI expert cautioned that sentiment analysis may not always capture human emotions accurately. Thus, psychological support provided via chatbots should be regarded as a complement to human interventions, which in our opinion can extend from healthcare professionals to coaches, teammates, friends, and family members.

### Nutrition

Chatbots have been used for nutrition advice ^17-19^. The nutritionist outlined multiple roles for chatbots in nutritional management, such as reminding athletes to stay hydrated, tracking dietary intake, and suggesting meal plans. A personalized dietary plan could employ an advanced AI algorithm to analyze factors like demographics, injury type, recovery stage, and allergy history, as well as signals from wearable devices and health tracking apps. The AI expert emphasized that building a personalized nutrition model demands a precise understanding of nutritional science and human physiology, as well as high-quality training data. However, given that chatbots might make mistakes such as recommending diets containing allergens ^20^ or harmful diet tips that promote eating disorders ^21^, they should be regarded as supplementary tools to human nutritionists rather than as their replacements.

### Tracking and Other Alternatives

Responses from the physiotherapist and the AI expert largely echoed those provided during the physical therapy round. The athlete noted that the automated tracking, recording, and reminding function helps reduce stress, echoing the psychologist’s comments. In line with remarks made by other researchers ^22^, the simulation highlighted several advantages of chatbots over traditional methods. These included reducing the need for manual reporting, offering convenient cloud-based access to records, real-time data collection, instantaneous analysis, and providing immediate advice. Despite these benefits, the simulation lacked a discussion on how chatbots could potentially enhance treatment outcomes over alternatives, such as increasing patient satisfaction or reducing recovery duration. In addition, the questions were designed to invoke engagements from all panel members. However, the nutritionist unexpectedly did not respond **(Table 3)**.

### Ethics

Distinct from other audience-initiated topics, questions regarding ethics prompted responses from all panelists **(Table 3)**. Some comments reiterated points from previous discussions, particularly regarding patient education. The conversation emphasized the crucial need for stringent adherence to medical privacy regulations such as HIPAA in the U.S. and GDPR in Europe. It highlighted the necessity of robust protocols for data encryption and storage to ensure security, as well as the needs for transparency on data collection, processing, and accessibility. However, the panel didn’t delve into the merits and drawbacks of open-source, locally deployed chatbots (especially those furnished with domain-specific knowledge) versus commercial and online chatbots with regards to privacy and security ^23^. Regarding bias and fairness, it was stressed that chatbot training should utilize diverse and representative datasets. As users, athletes should retain full discretion on whether to use chatbots, alternative methods, or a combination of both. The psychologist highlighted the need to implement chatbots in a manner that avoids triggering anxiety or other negative emotions. All the comments are in alignment with the five ethical principles proposed by AI4People: beneficence, non-maleficence, justice, autonomy, and explicability ^24^.

### Concluding question

The moderator was prompted to steer the panel discussion towards its end with a final question. As anticipated, the questions were all forward-thinking. Panelists offered predictions drawing from their respective fields of expertise. Foreseeing rapid advancements in AI and complementary technologies, the panel envisaged a future of precision sports rehabilitation in the chatbots era. In this vision, the rehabilitation program would be tailored to individual needs, bolstered by healthcare providers, and empowered by chatbots. According to responses from the simulated athlete, this form of personalized support would make rehabilitation feel like a natural part of the recovery process and the athlete taking charge of the rehabilitation journey.

## Discussions

The training data for ChatGPT spans nearly all disciplines, allowing it to role-play as an expert in response to specific inquiries. The interdisciplinary approach of PanelGPT brings several benefits. First, the responses come from panelists with complementary expertise, providing different perspectives that are automatically categorized. This aids in gaining a comprehensive view of the topic in question. For instance, including an athlete on the panel yielded a unique user perspective that could be overlooked in simple prompts, as demonstrated by the responses to “psychological support” (**Supplementary File 4**). Secondly, role-playing focuses the chatbot’s attention on the question and provides important contexts in responding to the questions. When the “physical therapy” questions were simply prompted to ChatGPT, the responses quickly drifted towards other topics like education and mental health (**Supplementary File 5**). Finally, having a panel of experts enables audiences to form a balanced view on a specific topic. For example, in addressing the “physical therapy” questions, the physiologist’s response highlighted the current limitations of chatbots in text or voice communication, while the AI expert expanded the discussion to the integration of real-time video analysis (**Supplementary Files 1 to 3**).

The breadth and depth of a response from a panelist depend on the training data set in the field. In several discussions, such as “patient education” and “tracking and other alternatives”, where we expected feedbacks from all panelists, there was a noticeable lack of direct responses from the nutritionist. It could be that the data set used to train ChatGPT for the nutritionist was under-representative in the rehabilitation field. Indeed, a combined search for ‘rehabilitation’ (or ‘rehab’) and ‘nutritionist’ (or ‘nutrition’) on PubMed yielded 6-to-8 times fewer hits compared to searches involving ‘physiotherapist’ (or ‘physiotherapy’) or ‘psychologist’ (or ‘psychology’) (as of 07/07/2023). To address this limitation, the human operator could send reminders to the nutritionist to elicit a response. The AI expert responded to questions in all topics. This is expected for the inherent need of AI expertise in the creation of such chatbot systems.

The data used to train ChatGPT only extends up until September 2021. As such, it was unable to provide comments that would reference more recent developments in chatbots like ChatGPT itself or BARD. The feature to activate BING within ChatGPT does allow for real-time information browsing from the internet. However, in practice this disrupted the panel discussion’s flow, resulting in a shift back to the regular ChatGPT conversation format and a subsequent loss of the expert identities after several exchanges (as shown in **Supplementary Files 6 to 8**).

We observed instances where the response to a question from the same expert was vague in one simulation but detailed in another. This suggests that conducting multiple simulations could enhance the efficacy of PanelGPT in providing a well-rounded understanding of the knowledge landscape surrounding an interdisciplinary topic. In fact, this practice enables self-consistency checking, which has been shown to improve reasoning performance of language models ^25^, and highlights recurring themes related to the question. Additionally, summarizing diverse responses from the multiple simulations facilitates the identification of contrasting viewpoints and emergent trends in the panel discussion.

Hallucination, the generation of unsupported or false information, is a prevalent issue with LLM-based chatbots. The multi-perspective approach of PanelGPT allows the chatbot to draw on the strengths and mitigate the weaknesses of each panelist when responding to specific questions. The current model is constrained by the fact that the same chatbot simulates all the panelists. With advances in chatbot development, this model could be extended by integrating responses from other LLM-chatbots, especially those possessing domain-specific knowledge. In this context, cross-referencing responses from different experts on the panel powered by distinct models help mitigate hallucination ^26^. Nonetheless, it remains crucial to cross-verify the conclusions drawn from the simulation with literature findings or opinions from human experts to ensure the accuracy of the information.

Throughout the simulation, we noted instances where comments from one expert were acknowledged by another. Intriguingly, contradictory comments between experts were not encountered. The richness and depth of the discussion can be further enhanced by utilizing additional prompting strategies. For instance, after each response round, panelists could be prompted to critically evaluate each other’s comments to foster consensus or highlight disagreements. Panelists may also be prompted to pose questions to one another, such as seeking clarifications or requesting further details on a given response. Moreover, panelists could be allowed to prompt the audience to clarify their questions if necessary. These additional prompting tactics not only make the panel discussion more engaging but also mirror a real-life scenario, increasing the likelihood of obtaining a thorough appreciation of the topic.

In conclusion, we present PanelGPT, an innovative method that capitalizes the multi-role-playing feature of ChatGPT to explore the knowledge landscape of interdisciplinary topics through simulated panel discussions. By applying this model to chatbots in sports rehabilitation, we summarized the opportunities and challenges in this emerging field. As a generalizable model, it could serve as a supplementary tool in the classroom, aiding students in understanding complex interdisciplinary topics. Furthermore, it can be extended to clinical settings to assist healthcare providers in enhancing patient care that requires interdisciplinary interventions, such as sports rehabilitation, stroke rehabilitation, and the management of recurrent pneumonia in long-term care facilities.

## Data Availability

Prompts and scripts for the final simulations are contained in the manuscript. Other data are available upon reasonable request to the authors.

## Data availability

Prompts and scripts from the simulations to support the conclusions are in Supplementary Files of the manuscript.

## Author Contributions

Conceptualization: G.H.; Formal data analysis: all authors; Writing - Original Draft: G.H; Writing - Review & Editing: all authors.

## Acknowledgements

NIH-NIGMS grants P20 GM103434, U54 GM-104942, and 1P20 GM121322 to GH; NSF 2125872 to GH and DAA; NIH-NLM grant No. R01LM013438 to LL; NIH-NLM grant 5R01LM013392 to DX. WVU Cancer Institute summer undergraduate research program to J.C.M. The content is solely the responsibility of the authors and does not necessarily represent the official views of the National Institutes of Health. The writing was polished by ChatGPT. We thank Zien Chen and Evelyn Shue from the lab for proof-reading.

## Supplementary File Legends

**Supplementary File 1:** Prompts and scripts for the 1^st^ round of simulation.

**Supplementary File 2:** Prompts and scripts for the 2^nd^ round of simulation.

**Supplementary File 3:** Prompts and scripts for the 3^rd^ round of simulation.

**Supplementary File 4:** Scripts for a direct prompt on “psychological support.”

**Supplementary File 5:** Scripts for a direct prompt on “physical therapy.”

**Supplementary File 6:** Prompts and scripts for the 1^st^ round of simulation with BING activated.

**Supplementary File 7:** Prompts and scripts for the 2^nd^ round of simulation with BING activated.

**Supplementary File 8:** Prompts and scripts for the 3^rd^ round of simulation with BING activated.

