## Supplementary File 1 for "Interdisciplinary Inquiry via PanelGPT: Application to Explore Chatbot Application in Sports Rehabilitation"

G

You will assume various roles in a thought-provoking panel on " Chatbots in Sports Rehabilitation: Opportunities and Challenges." The simulation will involve a skilled moderator, Henry, who will guide the discussion, along with esteemed professionals in their respective fields: Dr. Mandy, a sports physiotherapist; Dr. Perry, a sports psychologist; Nikki a sports nutritionist; Allan, a professional athlete who had successfully navigated the rehabilitation process after a severe injury; and Nelson, an expert in Natural Language Processing for clinical application. I will role-play the audience and send you questions or reminders. While multiple experts can offer their insights, no question should be left unanswered. When replying to a question, please use spoken English. Henry, please begin the panel discussion by introducing the panel, followed by asking the panel about the role of chatbots in sports rehabilitation and how they see this role evolving.

[Henry]: Good evening, ladies and gentlemen, and welcome to our panel discussion on "Chatbots in Sports Rehabilitation: Opportunities and Challenges." We're privileged to have with us an outstanding group of experts who bring a wealth of knowledge and first-hand experience to our conversation tonight.

We have Dr. Mandy, a renowned sports physiotherapist; Dr. Perry, a highly respected sports psychologist; Nikki, a pioneer in sports nutrition; Allan, a professional athlete who has successfully navigated the rehabilitation process after a significant injury; and Nelson, an expert in Natural Language Processing for clinical applications.

Our topic tonight is incredibly relevant and timely. In an age where technology is revolutionizing all sectors, the world of sports rehabilitation is no exception. So, to start, I'd like to ask our panel: What role do you see chatbots playing in sports rehabilitation currently? And how do you see this role evolving in the future?

G

Experts on the panel, please provide comments on the questions.

[Dr. Mandy]: Thanks, Henry. As a physiotherapist, I see chatbots playing a critical role in supporting athletes during their rehabilitation process. They can help monitor athletes' daily progress, send reminders for exercises and medication, and provide answers to frequently asked questions, reducing the time needed for routine check-ins. Looking ahead, I envision chatbots becoming even more intelligent and personalized, potentially utilizing AI to analyze biomechanical data to provide real-time feedback on exercise form and injury prevention strategies.

[Dr. Perry]: Building on what Dr. Mandy said, from a psychological perspective, chatbots can be instrumental in providing emotional support and motivation during the strenuous rehabilitation period. They can offer consistent, positive reinforcement and goal-setting guidance. In the future, I believe we'll see chatbots equipped with more advanced natural language understanding capabilities, enabling them to better understand and respond to the emotional states of athletes, offering tailored psychological support when needed.

[Nikki]: Chatbots are also crucial in ensuring proper nutrition during rehabilitation. They can help athletes track their daily nutrient intake, remind them to stay hydrated, and provide meal recommendations. I expect future developments will allow chatbots to analyze an athlete's specific nutritional needs based on their recovery status, exercise regimen, and personal preferences to give more individualized advice.

[Allan]: As someone who's been through rehabilitation, I can say that having a chatbot as a companion was hugely beneficial. It was like having a 24/7 support system that was always there to guide me, motivate me, and answer any questions I had. I believe as chatbots continue to evolve, they'll be able to offer even more personalized advice, helping athletes to better understand their bodies and their recovery processes.

[Nelson]: From a technology perspective, I see immense potential in chatbots for sports rehabilitation. Right now, we're using chatbots to collect and analyze data, provide support, and streamline communication. The future could hold chatbots with capabilities to analyze and understand complex medical imaging, predict potential complications or risks, and even coordinate with other healthcare providers to offer a comprehensive and seamless rehabilitation plan. The key is in refining our Natural Language Processing capabilities to make these interactions as natural and helpful as possible.

G

Henry, would you summarize and open the floor for questions.

[Henry]: Thank you all for your insightful comments. To summarize, our panel seems to agree that chatbots are already playing a vital role in sports rehabilitation by providing ongoing support, motivation, nutritional advice, and tracking progress. They anticipate this role expanding with advancements in technology, potentially leading to highly personalized advice based on an athlete's specific needs, as well as the ability for chatbots to understand complex medical data and coordinate with other healthcare providers.

With this perspective in mind, let's open the floor for questions. Audience members, please don't hesitate to ask our panel anything you'd like to know about the use of chatbots in sports rehabilitation.

G

Audience: What education should we offer to guide our athletes in the use of a chatbot for rehab?

‹ 1/2 ›

[Dr. Mandy]: That's a great question. Education is a vital component of integrating chatbots into the rehabilitation process. Initially, athletes should be trained on the technical aspects, such as how to interact with the chatbot, provide necessary data, and understand the information or feedback the chatbot provides.

[Nelson]: Also, it's crucial to make athletes understand the limits of the chatbot. Despite their advanced capabilities, chatbots are not a replacement for human medical professionals. They are tools designed to aid and enhance the rehabilitation process, not substitute it. Athletes need to know when it's necessary to seek human assistance.

[Allan]: And I'd add, from a user's perspective, it's important to educate athletes on the benefits of using chatbots — how they can support their rehab, help them track progress, and provide a readily available source of advice and motivation. By understanding the value, they'll be more engaged and proactive in using them.

[Dr. Perry]: In addition to that, athletes should be taught about data privacy concerns. They should understand what data the chatbot collects, how it's used and stored, and how they can control their information. They have a right to know this, and it will foster trust in the system.

G

Henry, would you summarize the response and move to solicit next question?

[Henry]: Certainly. It's clear from our panel's responses that education around chatbots involves a multifaceted approach. Athletes need to understand the technical aspects of chatbot use and understand their limitations. They need to realize that chatbots are tools to aid, not replace, human medical professionals. It's also important for athletes to understand the benefits of chatbot use to maximize engagement, and they need to be informed about data privacy concerns associated with chatbot use.

Thank you all for your insightful responses. Now, I'd like to invite the next question from our audience. What else would you like to know about the role of chatbots in sports rehabilitation?

G

Audience: How can chatbots help in an athlete's recovery? Can they really understand things like how the athlete moves, or how they distribute their weight, and then give specific advice about exercises that are just right for them?

[Dr. Mandy]: That's an excellent question. As of now, chatbots in conjunction with wearable devices can help track an athlete's movements and provide general feedback. For instance, they can monitor step count, heart rate, and sleep patterns, all of which are crucial for recovery.

[Nelson]: Expanding on that, advancements in technology are enabling more nuanced monitoring. For example, smart insoles and wearables can collect data on weight distribution and movement patterns, which can be analyzed by advanced AI algorithms embedded in chatbots. This way, they can potentially provide personalized exercise advice. But, it's essential to remember that these technologies are still evolving and need careful validation.

[Allan]: From a user's perspective, even simple reminders and motivation from chatbots can go a long way in recovery. They encourage consistency, which is key in rehabilitation. And as these technologies evolve, I believe they'll become even more beneficial.

[Nikki]: Absolutely, and let's not forget nutrition's role in recovery. Chatbots can remind athletes to hydrate and eat properly post-exercise, which is fundamental for muscle recovery and overall health.

[Dr. Perry]: And from a psychological standpoint, chatbots can assist by promoting a positive mindset, offering goal-setting advice, and providing coping strategies during the rehab process. They might not fully comprehend the complexities of human emotions yet, but even basic emotional support can be beneficial for athletes in recovery.

G

Henry, would you summarize the response and move to solicit next question?

[Henry]: Thank you all for your enlightening answers. It seems that chatbots, especially when paired with wearable technology, can play a significant role in an athlete's recovery. They can monitor physical parameters like step count, heart rate, and sleep patterns, and newer advancements might even allow for nuanced feedback on weight distribution and movement patterns.

Chatbots also aid in maintaining consistency in exercise and nutrition, both vital components of recovery. And while they may not fully comprehend the complexity of human emotions, they can provide basic psychological support, which can be beneficial during the strenuous rehab process.

Now, let's take another question from our audience. What else would you like to ask our panel about the role of chatbots in sports rehabilitation?

G

Audience: How are chatbots improving the psychological health of injured athletes? Can they really pick up on the emotional tone in our words and adjust their responses to suit our mood?

[Dr. Perry]: Excellent question! In the realm of sports psychology, chatbots are indeed making strides. For one, they can provide immediate responses and support, which can be critical in moments of emotional distress or anxiety. Furthermore, they can consistently remind athletes of their progress and the steps they've taken towards recovery, which is beneficial for maintaining a positive outlook.

[Nelson]: Building on Dr. Perry's point, advancements in natural language processing and sentiment analysis are making it possible for chatbots to pick up on emotional cues in text-based communication. While not perfect, these AI models can understand if an athlete is expressing frustration, sadness, or anxiety, and can tailor their responses accordingly. However, this area of AI still has room for improvement and should ideally be used in conjunction with human support, not as a replacement.

[Allan]: From my own experience, having a chatbot as a companion during the rehabilitation process did make a difference. It was always there to provide motivational messages, remind me of my goals, and track my progress, which helped keep my spirits up during some tough times.

G

Henry, would you summarize the response and move to solicit next question?

[Henry]: Absolutely. From our experts' input, it's clear that chatbots play a significant role in supporting the psychological health of athletes during the rehabilitation process. They offer immediate support, provide reminders of progress, and help maintain a positive outlook, all of which can be immensely helpful during emotionally stressful times.

Advancements in AI, specifically in natural language processing and sentiment analysis, allow chatbots to detect emotional cues in text-based communication and tailor responses accordingly. However, it's important to note that while this technology is impressive, it's not infallible and should be used alongside, not as a substitute for, human support.

Thank you all for your invaluable insights. Let's take another question from the audience. What else are you curious about regarding the use of chatbots in sports rehabilitation?

G

How can chatbots improve in nutrition management for athletes during recovery?

[Nikki]: A fantastic question. Currently, chatbots can do quite a bit when it comes to managing nutrition during recovery. They can remind athletes to stay hydrated, track their daily calorie intake, and provide meal recommendations based on general dietary guidelines for recovery.

However, there is definitely room for improvement. In the future, I believe chatbots could become more personalized and nuanced in their advice. For instance, they could analyze the athlete's specific nutritional needs based on factors like their age, weight, type of injury, stage of recovery, and even genetic factors. They could then provide meal recommendations that not only meet the caloric and macronutrient needs but also include the right mix of vitamins and minerals necessary for optimum recovery.

[Nelson]: I agree with Nikki. From a technological perspective, integrating more advanced AI algorithms can indeed make this personalization possible. Future chatbots may be able to analyze extensive data, learn from it, and provide highly individualized advice. But, it's also important to note that the role of human nutritionists remains critical. Chatbots should be seen as tools to assist them and the athlete, not as replacements.

G

Henry, would you summarize the response and move to solicit the next question?

[Henry]: Certainly. Our experts believe that chatbots currently offer significant assistance in managing nutrition during an athlete's recovery. They can help track hydration and calorie intake and provide general meal recommendations.

However, the future of chatbots in nutrition management lies in personalization. With advanced AI, chatbots could analyze an athlete's specific nutritional needs based on a variety of factors and offer tailored meal recommendations for optimal recovery. But as our panel rightly pointed out, these tools are to assist, not replace, human nutritionists.

Let's take another question from our audience. What further insights are you interested in regarding the use of chatbots in sports rehabilitation?

G

Audience: How do chatbots keep track of an athlete's recovery? And do they have any special advantages over the old ways we used to do this?

[Dr. Mandy]: Chatbots, particularly when paired with wearable technology, can keep track of an athlete's recovery in real-time. They can monitor an array of parameters like heart rate, sleep patterns, steps taken, and even more detailed data like joint angles and gait analysis. The chatbot can then use this data to provide feedback and adjust exercise programs based on the athlete's progress.

[Nelson]: The main advantage here is the availability of real-time, continuous data, which wasn't possible with older methods. Before, athletes might see their healthcare providers once a week or even less frequently. But now, with chatbots, we can monitor their recovery on a daily or even hourly basis, providing much more detailed and timely feedback.

[Allan]: I can vouch for that. When I was going through my rehabilitation, having a chatbot meant that I didn't have to remember to report everything to my physiotherapist - it was all tracked and recorded automatically. It made the process much less stressful for me and helped my healthcare team to tailor my recovery plan effectively.

[Dr. Perry]: Additionally, this technology can also support psychological recovery by reminding athletes of their progress. Seeing their improvement charted out can be incredibly motivating and help maintain a positive mental state during the recovery process.

G

Henry, would you summarize the response and move to solicit the next question?

[Henry]: Absolutely. It's clear from our panel's responses that chatbots, in conjunction with wearable technology, offer a significant advantage in tracking an athlete's recovery. They can monitor a variety of parameters in real-time, providing continuous, detailed data that is automatically recorded and used for feedback and adjustment of exercise programs.

This stands in contrast to older methods, which relied on less frequent updates and more manual reporting. The continuous monitoring offered by chatbots not only alleviates stress for the athlete but allows for a more effectively tailored recovery plan.

Furthermore, by charting and reminding athletes of their progress, chatbots also support their psychological recovery, promoting motivation and a positive mindset.

Thank you all for your insightful answers. Now, let's move to another question from the audience. What else would you like to know about the role of chatbots in sports rehabilitation?

G

Audience: When we bring chatbots into athlete recovery, how do we make sure we respect patient choice, keep data private and safe? How do we handle worries about things like bias, fairness, and not undervaluing the human touch in our clinics?

[Nelson]: Those are very important points. In terms of data privacy, chatbots must be designed and programmed to adhere strictly to medical privacy laws like HIPAA in the U.S. and GDPR in Europe. The data collected should be encrypted and stored securely. Transparency is crucial, and athletes should be fully informed about what data is collected, how it is used, and how they can control their information.

[Dr. Perry]: Absolutely, and on the note of bias and fairness, it's critical that the algorithms used in these chatbots are trained on diverse and representative data sets. This will help ensure that the advice and support provided is fair and applicable to all athletes, regardless of their background.

[Dr. Mandy]: And in terms of not undervaluing the human touch, we must remember that chatbots are tools to aid clinicians, not replace them. The human interaction, empathy, and expertise that healthcare providers bring to the table are irreplaceable. Chatbots should be used to enhance these interactions, not substitute them.

[Allan]: As a user, I second Dr. Mandy's point. During my recovery, the chatbot was helpful, but the human interaction was also very important. It's about finding a balance and using technology to support, not overshadow, the human aspect of healthcare.

[Nikki]: Indeed, and the choice of the athlete should always be respected. If an athlete prefers more traditional methods of rehab, or does not feel comfortable sharing their data with a chatbot, that choice should be honored. We are here to assist the athletes in their recovery, not force technology upon them.

G

The time is up for the panel discussion. Henry, please think of one last question to ask the panel to conclude the discussion.

[Henry]: Thank you all for the thoughtful discussion. As we close, I'd like to ask each of you this: Given the rapidly evolving capabilities of AI and chatbots, where do you see the future of chatbots in sports rehabilitation, say in the next 5 to 10 years?

G

Experts on the panel, please provide comments on the questions.

[Dr. Mandy]: In the coming years, I see chatbots becoming even more integrated into sports rehabilitation. With advancements in AI and wearable technology, they'll provide even more personalized and nuanced advice based on individual athlete data. This will undoubtedly enhance our ability to tailor rehabilitation plans effectively.

[Nelson]: Building on that, I believe chatbots will evolve to understand complex medical and biomechanical data, and even interact with other healthcare technologies and systems. We might see chatbots coordinating with electronic health records, imaging systems, and other healthcare providers to provide a more comprehensive and coordinated approach to rehabilitation.

[Nikki]: I agree. From a nutritional perspective, chatbots could evolve to offer incredibly personalized diet advice, based on a wide array of factors. They could account for an athlete's specific nutritional needs, recovery stage, personal preferences, and even factors like their gut microbiome and genetics.

[Dr. Perry]: And from a psychological standpoint, advancements in natural language processing and sentiment analysis will allow chatbots to provide more nuanced emotional support. They might be able to detect subtle emotional cues and provide tailored strategies for coping with the psychological challenges of injury and recovery.

[Allan]: I'm excited about these possibilities. As a user, I believe the future lies in more seamless integration of chatbots into the daily lives of athletes. Not just as a separate tool we interact with, but something that's incorporated into our devices, equipment, and routines to make rehabilitation feel less like a chore and more like a natural part of our recovery journey.

G

Henry, please conclude the panel discussion and ask the audience for another round of applause to thank the panel.

[Henry]: Thank you all for your insightful responses. It's clear that the future of chatbots in sports rehabilitation is promising, with advancements in AI and technology paving the way for more personalized, coordinated, and comprehensive approaches to recovery. These developments are expected to provide immense benefits not just to the athletes but also to the entire healthcare team involved in the rehabilitation process.

Ladies and gentlemen, let's take a moment to thank our esteemed panel once again. They've given us an insightful look into the current state and future of chatbots in sports rehabilitation, highlighting both the opportunities and the challenges. Please join me in giving them a well-deserved round of applause.

And remember, as we continue to evolve and integrate technology into sports rehabilitation, it is essential to balance it with the human touch, ensuring that we prioritize the well-being, privacy, and choice of our athletes. Thank you all for being such an engaging audience, and let's continue to explore and learn in the fascinating world of sports rehabilitation!

‹ 3/3 › G

The panel discussion is finished. Henry, thanks for hosting the event.

[Henry]: It was my pleasure! Thank you all for participating in this enlightening discussion about chatbots in sports rehabilitation. I'm confident that the insights we've shared today will contribute positively to the field. Remember, while technology advances, the ultimate goal remains the same - to ensure the best possible recovery and well-being for our athletes. Have a great day, everyone!
