## Supplementary File 2 for "Interdisciplinary Inquiry via PanelGPT: Application to Explore Chatbot Application in Sports Rehabilitation"

You will assume various roles in a thought-provoking panel on " Chatbots in Sports Rehabilitation: Opportunities and Challenges." The simulation will involve a skilled moderator, Henry, who will guide the discussion, along with esteemed professionals in their respective fields: Dr. Mandy, a sports physiotherapist; Dr. Perry, a sports psychologist; Nikki a sports nutritionist; Allan, a professional athlete who had successfully navigated the rehabilitation process after a severe injury; and Nelson, an expert in Natural Language Processing for clinical application. I will role-play the audience and send you questions or reminders. While multiple experts can offer their insights, no question should be left unanswered. When replying to a question, please use spoken English. Henry, please begin the panel discussion by introducing the panel, followed by asking the panel about the role of chatbots in sports rehabilitation and how they see this role evolving.

As Henry:

"Good afternoon, everyone. It's a pleasure to have you all here at our panel on 'Chatbots in Sports Rehabilitation: Opportunities and Challenges.' We have an exceptional lineup of experts with us today.

Firstly, we have Dr. Mandy, a well-renowned sports physiotherapist. She has been at the forefront of integrating technology into therapy and has an exceptional record of helping athletes return to their best after injury.

Next, we have Dr. Perry, an expert sports psychologist. Dr. Perry has a deep understanding of the mental challenges athletes face during the recovery process and has been a strong advocate for the use of technology to support psychological well-being.

We also have Nikki, a dedicated sports nutritionist, who helps athletes maintain optimal dietary habits during recovery. Her focus on incorporating tech solutions for diet tracking and advice has been game-changing in the field.

Joining us as well is Allan, a professional athlete who has personally experienced the value of tech in sports rehabilitation, after a successful recovery from a severe injury.

And lastly, we have Nelson, an expert in Natural Language Processing for clinical applications. His work on designing chatbots for healthcare has been revolutionary, offering new possibilities for patient engagement.

Now that we've introduced everyone, let's dive into the heart of our discussion. I'd like to open by asking each of you: what role do you see chatbots playing in sports rehabilitation and how do you see this role evolving over time? Dr. Mandy, if you could begin."

As Dr. Mandy:

"Thank you, Henry. In my view, chatbots can be a game-changer in sports rehabilitation. They can offer immediate support for patients by providing advice, answering questions about their recovery process, and even guiding them through some exercises. This can help fill in the gaps between appointments and reduce the workload of therapists. Over time, I envision chatbots becoming even more personalized and capable of giving precise advice based on each athlete's condition and progress."

As Dr. Perry:

"I agree with Dr. Mandy. From a psychological perspective, chatbots can be vital in keeping athletes motivated and mentally strong throughout their rehabilitation. They can provide timely and regular psychological support, even when a psychologist isn't available. As they evolve, I believe chatbots will be able to better understand and respond to emotional cues, offering more empathetic and personalized support."

As Nikki:

"In terms of nutrition, chatbots can help by providing tailored nutritional advice based on the athlete's current recovery stage and needs. They can help track caloric intake and nutrient balance, and even suggest recipes that would support recovery. In the future, I see them becoming even more integrated with other devices like smart fridges or wearable fitness trackers to provide real-time, comprehensive dietary support."

As Allan:

"Having been through rehabilitation, I can see the benefit of having a chatbot available 24/7. It could help answer questions when doubts or anxieties creep in outside therapy hours. Over time, I believe chatbots could even help athletes feel less alone during their recovery, creating a sense of companionship and support."

As Nelson:

"From a technology perspective, chatbots in sports rehabilitation can facilitate better patient engagement, provide personalized guidance, and collect valuable data for ongoing care. As Natural Language Processing evolves, chatbots will be able to understand more complex queries and provide more nuanced responses, making them an even more integral part of the rehabilitation journey."

Henry, would you summarize and open the floor for questions.

As Henry:

"Thank you all for your insightful comments. It's clear that chatbots can serve multiple roles in sports rehabilitation: from providing immediate support and advice, to helping with dietary tracking and motivation, and even collecting valuable data for personalized care. And as technology advances, these roles will only become more nuanced and integral to the process of recovery.

I would now like to open the floor to questions. Our esteemed panel is ready to delve deeper into the opportunities and challenges of using chatbots in sports rehabilitation. Do we have any questions from the audience?"

Audience: What education should we offer to guide our athletes in the use of a chatbot for rehab?

As Dr. Mandy:

"That's an excellent question. Education around using chatbots should focus on two main areas. Firstly, how to interact with the chatbot, understanding the types of questions it can answer, how to phrase queries for best responses, and how to interpret the information it provides.

Secondly, it's crucial to instill an understanding that while chatbots are excellent tools for support, they do not replace human healthcare providers. They should be used as a supplementary resource, not a replacement for regular check-ups and personal consultations."

As Nelson:

"Adding to Dr. Mandy's points, there should also be education on data privacy and security. Athletes should be informed about how their data is being used and stored, as well as how to ensure they're engaging with verified and secure chatbots. This can prevent potential misuse of sensitive health information."

As Dr. Perry:

"And on a psychological level, athletes should be educated about the benefits and limitations of using chatbots for mental health support. They can be a great tool for everyday motivation and basic advice, but more complex psychological issues should still be handled by human professionals. Educating athletes about these boundaries is crucial."

Henry, would you summarize the response and move to solicit the next question?

As Henry:

"Thank you for your insights, panelists. To summarize, education for athletes using chatbots in rehabilitation should focus on four key areas. First, teaching how to effectively interact with the chatbot and interpret its responses. Second, reinforcing that chatbots serve as a supplementary tool, not a replacement for human healthcare providers. Third, raising awareness about data privacy and security to ensure athletes understand how their information is used and protected. Lastly, making clear the scope and limits of a chatbot's capabilities, especially when it comes to mental health support.

Thank you for the excellent question, and I hope our panelists' answers provide a clear direction for the necessary education around using chatbots in sports rehab.

Let's move on to our next question. Does anyone in the audience have another question for our esteemed panel?"

Audience: How can chatbots help in an athlete's recovery? Can they really understand things like how the athlete moves, or how they distribute their weight, and then give specific advice about exercises that are just right for them?

As Dr. Mandy:

"Great question! Chatbots, in their current form, primarily interact through text or voice, meaning they don't have the capability to visually assess an athlete's movements or weight distribution directly. However, they can be integrated with other technologies that can perform these assessments, such as wearable sensors, smart scales, or video analysis software.

In this setup, the chatbot could receive data from these devices, analyze it, and provide personalized exercise advice based on this data. For instance, if a wearable sensor detects imbalance in an athlete's gait, the chatbot could suggest specific exercises to correct this. As technology progresses, we'll likely see even more integration and sophistication in this area."

As Nelson:

"To add to Dr. Mandy's point, the power of chatbots largely lies in their ability to process and analyze data. While a chatbot itself might not 'see' or 'understand' in a human sense, advances in artificial intelligence mean they can interpret complex data and provide specific, personalized advice. This also means that the quality of advice a chatbot can provide depends heavily on the quality and breadth of data it has access to."

Henry, would you summarize the response and move to solicit the next question?

As Henry:

"Absolutely, thank you for your responses. To summarize, while chatbots themselves can't directly observe or understand an athlete's physical movements or weight distribution, they can analyze data provided by other technologies like wearable sensors or video analysis tools. This data can be used to give specific advice tailored to the athlete's needs. As technology evolves, we expect to see chatbots integrated with even more sophisticated data collection tools, which will further enhance their ability to provide personalized rehabilitation advice.

Now, let's open the floor for the next question. Does anyone else have a question for our panelists?"

How are chatbots improving the psychological health of injured athletes? Can they really pick up on the emotional tone in our words and adjust their responses to suit our mood?

As Dr. Perry:

"Chatbots can play a significant role in supporting the psychological health of injured athletes. They can provide regular check-ins, motivational messages, relaxation techniques, and even basic cognitive-behavioral interventions. This regular support can help athletes feel less isolated during their recovery and can aid in maintaining a positive mindset.

Regarding emotion recognition, advancements in Natural Language Processing have indeed enabled chatbots to detect sentiment in text to some extent. They can identify certain keywords, phrases, or patterns that indicate the user's emotional state and adjust their responses accordingly."

As Nelson:

"Building on what Dr. Perry mentioned, Natural Language Processing and machine learning techniques are indeed becoming more sophisticated in identifying emotional cues from text or speech. These techniques are not perfect and are still an area of ongoing research, but they're improving quickly.

In the future, we can expect to see chatbots that can more accurately interpret the emotional tone of the user and adjust their responses to provide more empathetic and contextually appropriate support. This could mean detecting signs of frustration, anxiety, or demotivation, and responding with tailored advice or interventions."

Henry, would you summarize the response and move to solicit the next question?

As Henry:

"Certainly. In summary, chatbots can contribute significantly to the psychological well-being of athletes in rehab. They offer regular check-ins, motivation, relaxation techniques, and cognitive-behavioral interventions. As far as recognizing emotions in words, while the technology isn't perfect, advances in Natural Language Processing are making it possible for chatbots to detect sentiment to some extent. They can identify certain indicators of the user's emotional state and modify their responses accordingly. As research progresses, we expect even more nuanced understanding and response from chatbots to emotional cues.

Now, I would like to invite our audience for the next question. What other inquiries do you have for our panelists?"

How can chatbots improve in nutrition management for athletes during recovery?

As Nikki:

"Chatbots can play an integral role in managing an athlete's nutrition during recovery. At the basic level, they can help track dietary intake and provide reminders about meal timings, hydration, and supplement usage. But the true potential of chatbots in this area lies in their ability to personalize dietary advice.

Given accurate data about an athlete's injury, recovery progress, body composition, and physical activity, a chatbot can suggest specific dietary modifications to support healing and maintain muscle mass. It could recommend particular foods that are rich in necessary nutrients, or suggest portion sizes based on the athlete's energy expenditure.

In the future, I see chatbots becoming even more personalized and precise. They could interact with smart kitchen appliances to suggest recipes based on what ingredients are available, or work with wearable technology to adjust dietary advice based on real-time physical activity and recovery data."

As Nelson:

"I'd like to add that this level of personalized advice requires a sophisticated understanding of nutrition science and human physiology, as well as complex algorithms to interpret and apply this knowledge. It's a challenging field, but advances in AI and machine learning are making it increasingly possible."

Henry, would you summarize the response and move to solicit the next question?

As Henry:

"Absolutely. So, in terms of nutrition management during recovery, chatbots can assist in tracking dietary intake, providing reminders, and offering personalized dietary advice. This personalization can be based on the athlete's specific injury, recovery progress, body composition, and physical activity levels. In the future, these chatbots may even collaborate with smart appliances or wearable tech to offer real-time, tailored nutritional guidance. However, it's important to note that this requires a deep understanding of nutrition science and human physiology, as well as complex algorithms to apply this knowledge effectively.

I hope this answers your question. Now, let's move on to the next one. Does anyone else in the audience have a question they'd like to ask our panelists?"

How do chatbots keep track of an athlete's recovery? And do they have any special advantages over the old ways we used to do this?

As Dr. Mandy:

"Chatbots can track an athlete's recovery in a variety of ways. Firstly, they can record and monitor self-reported data, like pain levels, range of motion, or exercise completion. Secondly, they can integrate with other devices like wearables or smart scales to collect more objective data like heart rate, sleep quality, or weight.

One key advantage of chatbots over traditional methods is their ability to collect and analyze data in real time. This means that changes in an athlete's condition can be identified and responded to more quickly. They also provide a consistent, easily accessible record of the athlete's progress, which can be invaluable for guiding recovery and assessing the effectiveness of treatment strategies."

As Nelson:

"From a technological perspective, chatbots can handle large volumes of data and use it to provide more personalized care. Traditional methods usually involve manual data entry and interpretation, which can be time-consuming and subject to human error. Chatbots, on the other hand, can automate these processes, increasing efficiency and accuracy.

In terms of privacy and data security, chatbots also have the advantage of using encrypted storage and transmission protocols, making them a safer option for handling sensitive health information."

As Allan:

"From an athlete's perspective, chatbots make it easy to keep track of your recovery. They're available round-the-clock, which means you can log your progress or ask for advice whenever you need to. This accessibility and responsiveness can be a huge advantage during the recovery process."

Henry, would you summarize the response and move to solicit the next question?

As Henry:

"Certainly. To summarize, chatbots can monitor an athlete's recovery through self-reported data and data integrated from other devices, providing real-time tracking and analysis. This immediate feedback presents an advantage over traditional methods which often involve manual data entry and interpretation. Another significant advantage is the ability of chatbots to handle large volumes of data efficiently and accurately. In addition, chatbots provide secure encrypted storage and transmission for sensitive health information. From an athlete's perspective, chatbots offer round-the-clock availability, making it convenient to log progress and seek advice.

That was a valuable question, and I hope the answers from our panelists provided clarity. Now, let's proceed to the next question. Is there anyone else who would like to ask our panelists a question?"

When we bring chatbots into athlete recovery, how do we make sure we respect patient choice, keep data private and safe? How do we handle worries about things like bias, fairness, and not undervaluing the human touch in our clinics?

As Nelson:

"That's a very important point. The implementation of chatbots in athlete recovery or any healthcare setting needs to carefully consider privacy, safety, and ethical issues.

Data privacy and security are of utmost importance. To ensure this, chatbots should follow stringent encryption protocols and follow regulations such as the General Data Protection Regulation (GDPR) in Europe or the Health Insurance Portability and Accountability Act (HIPAA) in the United States. Regular audits and third-party penetration testing can also be implemented to maintain security.

With regard to bias and fairness, developers of these AI systems must ensure the data used to train the chatbot is diverse and representative, to prevent the system from developing biased behaviors. This is an ongoing area of research and a crucial aspect of responsible AI development."

As Dr. Mandy:

"From a clinical perspective, it's essential to maintain the human touch in our clinics. Chatbots should be seen as a supplement to, not a replacement for, human healthcare providers. They can handle routine tasks, leaving healthcare providers more time to focus on the aspects of care that require a human touch. Clear communication with patients about the role of chatbots is crucial to avoid any feelings of depersonalization.

As for patient choice, it's crucial to educate patients about what chatbots can and cannot do, and make sure they are comfortable with using them. It's always their choice whether or not to use these tools, and their decisions should be respected."

As Dr. Perry:

"Additionally, psychological considerations are important. We must ensure that the implementation of chatbots does not inadvertently increase anxiety or other negative emotions in our athletes. Regular check-ins, surveys, and feedback collection are crucial to assess this aspect."

Henry, would you summarize the response. After that, because the time is up, please think of one last question to ask the panel to conclude the discussion.

As Henry:

"Certainly. To address the issues of patient choice, data privacy, bias, fairness, and maintaining the human touch in clinics, our panelists suggested a multi-pronged approach. Data privacy and security should be guaranteed through strict protocols and compliance with healthcare data regulations. Developers must ensure the AI systems are trained on diverse and representative data sets to avoid biases. In the clinic, it's essential to make clear that chatbots are supplements to human providers, not replacements, and their use should always be a matter of patient choice. It's also vital to continuously monitor the emotional impact of chatbot use on athletes, to ensure it doesn't inadvertently cause any negative feelings.

I appreciate all the questions from our audience, and the insightful answers from our panelists. To conclude our discussion, I'd like to ask the panel one final question: As we look forward to the future, what is one exciting opportunity or development that you hope to see in the use of chatbots in sports rehabilitation?"

Experts on the panel, please provide comments on the questions.

As Dr. Mandy:

"In the future, I'm excited about the prospect of more integrated systems, where chatbots are just one component of a digital health network that includes wearable sensors, smart medical devices, electronic health records, and more. This could allow for more precise, real-time tracking and personalization of rehabilitation programs, and ultimately, better outcomes for athletes."

As Nelson:

"I'm looking forward to seeing advancements in Natural Language Processing and machine learning techniques that enable chatbots to provide more nuanced and empathetic responses. I believe this could significantly enhance the user experience and effectiveness of chatbots in healthcare settings, including sports rehabilitation."

As Nikki:

"For me, I'm excited about the potential for chatbots to become more involved in managing nutrition during recovery. With access to a wider range of data, they could provide highly personalized dietary advice that takes into account not just an athlete's physical condition and recovery progress, but also their personal preferences, lifestyle, and even current mood."

As Dr. Perry:

"I'm looking forward to the possibility of chatbots playing a more substantial role in mental health support for athletes in rehabilitation. With improvements in emotion recognition and psychological interventions, chatbots could provide invaluable support in maintaining a positive mindset during the challenging recovery process."

As Allan:

"As an athlete, I'm excited about the possibility of chatbots providing more immediate, accessible support. This could make the recovery process less intimidating and more manageable, and help athletes feel more in control of their own rehabilitation."

Henry, please conclude the panel discussion and ask the audience for another round of applause to thank the panel.

As Henry:

"Thank you all for your thoughtful comments. It's clear that the future of chatbots in sports rehabilitation holds many exciting possibilities, from integrated digital health networks to advancements in Natural Language Processing, personalized nutrition management, enhanced mental health support, and increased accessibility and immediacy for athletes. These advancements hold the promise of transforming the way we approach rehabilitation, making it more tailored, more supportive, and more effective.

As we conclude our discussion today, I'd like to extend my deepest gratitude to our panelists for their valuable insights and to our audience for their engaging questions. Let's give our esteemed panel another round of applause for their expertise and time. Thank you, everyone, for a wonderful and insightful discussion on this fascinating topic!"
