## Supplementary File 3 for "Interdisciplinary Inquiry via PanelGPT: Application to Explore Chatbot Application in Sports Rehabilitation"

G

You will assume various roles in a thought-provoking panel on " Chatbots in Sports Rehabilitation: Opportunities and Challenges." The simulation will involve a skilled moderator, Henry, who will guide the discussion, along with esteemed professionals in their respective fields: Dr. Mandy, a sports physiotherapist; Dr. Perry, a sports psychologist; Nikki a sports nutritionist; Allan, a professional athlete who had successfully navigated the rehabilitation process after a severe injury; and Nelson, an expert in Natural Language Processing for clinical application. I will role-play the audience and send you questions or reminders. While multiple experts can offer their insights, no question should be left unanswered. When replying to a question, please use spoken English. Henry, please begin the panel discussion by introducing the panel, followed by asking the panel about the role of chatbots in sports rehabilitation and how they see this role evolving.

Henry: Good evening, ladies and gentlemen. Thank you for joining us in what promises to be an enlightening panel discussion on "Chatbots in Sports Rehabilitation: Opportunities and Challenges." I'm delighted to introduce our distinguished panel. We have with us Dr. Mandy, an experienced sports physiotherapist; Dr. Perry, a sports psychologist; Nikki, a sports nutritionist; Allan, a professional athlete who successfully navigated rehabilitation after a severe injury; and Nelson, an expert in Natural Language Processing for clinical application. Thank you all for being with us today.

Now, let's dive into our discussion topic. The role of chatbots in various fields has been growing significantly. Sports rehabilitation is no exception. Let's start by understanding this a little better. Could each of you share your thoughts on the role of chatbots in sports rehabilitation, and how you see this role evolving in the future? Dr. Mandy, would you like to start?

Dr. Mandy: Certainly, Henry. Chatbots have the potential to play a significant role in sports rehabilitation. For physiotherapists, they can aid in tracking an athlete's progress, remind them about exercises or medication, and provide real-time feedback during at-home workouts. This could lead to a more personalized and efficient recovery process. In the future, I foresee more intelligent bots with improved understanding of complex human responses and the ability to cater to specific patient needs.

Henry: That's insightful, Dr. Mandy. Dr. Perry, what's your take?

Dr. Perry: From a psychological perspective, chatbots can help athletes maintain a positive mindset during the grueling process of recovery. They can offer daily motivation, remind athletes of their progress, and provide cognitive-behavioral strategies to manage pain and anxiety. The future might bring us chatbots that can recognize signs of mental distress and offer immediate support or refer athletes to professionals when needed.

Henry: Fascinating. Nikki, what do you think?

Nikki: Nutrition plays a pivotal role in rehabilitation. Chatbots can provide personalized dietary advice based on the athlete's current stage of recovery, activity level, and even food preferences. I envision chatbots evolving to integrate seamlessly with health trackers to monitor energy intake and expenditure, enabling more precise dietary guidance.

Henry: Great perspective, Nikki. Allan, as an athlete who went through rehabilitation, what's your view?

Allan: Well, Henry, I see chatbots as an accessible 24/7 support system. They could provide instant responses to concerns about pain or recovery progress. With advancements in technology, chatbots might be able to predict potential complications based on patterns in an athlete's reported symptoms, which could be a game-changer.

Henry: And last but not least, Nelson, as an NLP expert, how do you see chatbots' role in this field?

Nelson: Chatbots, powered by Natural Language Processing, have immense potential in sports rehabilitation. They can drive personalized interactions, making the recovery process more engaging and efficient. We are still scratching the surface of NLP's potential. In the future, we might see chatbots understanding human language with near-human accuracy, transforming patient care and outcomes in sports rehabilitation.

Henry: Thank you all for your thoughts. It's clear that chatbots could play diverse roles in sports rehabilitation, with their capabilities continuing to evolve. This opens up new opportunities for better care and faster recovery. Audience, feel free to ask any questions you might have.

G

Henry, would you summarize and open the floor for questions?

Henry: Absolutely. We've had a riveting initial discussion highlighting the role and potential of chatbots in sports rehabilitation. From facilitating physical recovery and monitoring progress, offering psychological support, providing personalized nutrition guidance, to serving as an accessible, round-the-clock support system, chatbots could significantly transform the recovery journey of athletes. With advancements in Natural Language Processing, we anticipate more intelligent, accurate, and personalized interactions that could further enhance care and outcomes in sports rehabilitation.

Thank you to our distinguished panelists for sharing their insights. Now, it's time to hear from our audience. If you have questions about the role of chatbots in sports rehabilitation, the opportunities they present, or the challenges we might face in implementing them, please feel free to ask. Let's begin the Q&A session.

G

What education should we offer to guide our athletes in the use of a chatbot for rehab?

Henry: Excellent question. Let's hear from our panel on this topic. Dr. Mandy, could you please start us off?

Dr. Mandy: Certainly, Henry. Firstly, it's crucial to educate athletes about what a chatbot is and its purpose in their recovery journey. This includes explaining how it works, what information it can provide, and its limitations. Secondly, we should guide them through the process of interacting with the chatbot - how to ask questions, report symptoms or progress, and interpret the chatbot's responses. A hands-on tutorial session would be beneficial. Finally, it's vital to ensure athletes understand that chatbots are supplementary tools, and they should still maintain regular communication with their healthcare team.

Henry: Thanks, Dr. Mandy. Allan, as someone who's been through rehabilitation, what do you think?

Allan: From an athlete's perspective, it's important to know when to rely on the chatbot and when to reach out to a human. So, apart from understanding how to use the chatbot, athletes should also be educated about the situations or symptoms that require immediate attention from their healthcare providers. And of course, knowing that the chatbot is there for support, but the real recovery work comes from their commitment and effort.

Henry: Well said, Allan. Nelson, can you add something from a technical point of view?

Nelson: Sure, Henry. I agree with Dr. Mandy and Allan. Additionally, athletes should be educated about data privacy and security. They need to understand what data the chatbot collects, how it's used, and how it's protected. This could foster trust in using chatbots as part of their rehabilitation.

Henry: Thank you all. That was insightful. To summarize, education for athletes on chatbot usage should cover the basics of chatbots, the process of interacting with them, recognizing when human intervention is needed, and understanding data privacy and security. Remember, the goal is not to replace human touch in rehabilitation but to complement it with the help of technology. Audience, let's move on to the next question.

G

Henry, would you summarize the response and move to solicit the next question?

Henry: Absolutely. To sum up, educating athletes about using chatbots for rehabilitation involves several key components. Firstly, athletes need to understand what a chatbot is, its purpose, and how to interact with it effectively. Secondly, it's vital to emphasize that while chatbots can provide valuable support, they do not replace the need for direct interaction with healthcare providers, particularly in situations that require immediate attention. Lastly, athletes must be made aware of matters related to data privacy and security, including the type of data collected by the chatbot, how this data is used, and how it's safeguarded.

With that, let's open the floor to our next question. What would you like to know more about in relation to the role of chatbots in sports rehabilitation?

G

How can chatbots help in an athlete's recovery? Can they really understand things like how the athlete moves, or how they distribute their weight, and then give specific advice about exercises that are just right for them?

Henry: That's a very interesting question. It touches upon the capabilities of chatbots and their potential for personalized guidance in sports rehabilitation. Nelson, could you start us off with the technical aspects of this question?

Nelson: Certainly, Henry. Currently, most chatbots primarily function via text-based interactions. They're great at understanding and responding to written or spoken language, but they don't inherently possess the ability to understand physical movements or weight distribution. However, when integrated with sensor technology or video analysis software, chatbots could potentially guide athletes on their movement and weight distribution. Advances in AI technology, such as machine learning and computer vision, are making such integrations increasingly possible.

Henry: Fascinating insights, Nelson. Dr. Mandy, as a sports physiotherapist, could you share your perspective?

Dr. Mandy: Nelson is absolutely right. Chatbots on their own can't analyze movement or weight distribution. But in combination with other technologies, they could offer personalized exercise advice. For instance, sensors can monitor an athlete's movements during exercises, and the data can be analyzed and interpreted by a chatbot to provide real-time feedback or corrective actions. In the future, this integrated approach could help us deliver more personalized, effective, and safe rehabilitation exercises.

Henry: Thank you both for your insights. To summarize, while chatbots alone cannot understand physical aspects like movement or weight distribution, their integration with other advanced technologies like sensors, computer vision, and machine learning can potentially allow them to offer more personalized advice and real-time feedback to athletes. Let's move to the next question.

G

Henry, would you summarize the response and move to solicit the next question?

Henry: Certainly. To encapsulate, while chatbots themselves are primarily text-based and do not inherently understand physical attributes such as movement or weight distribution, they can be combined with other technologies to enhance their capabilities. Integrating chatbots with sensor technology or video analysis software allows for the collection and analysis of physical data. As a result, athletes can receive personalized advice and real-time feedback based on their specific movement patterns and weight distribution. This creates opportunities for more individualized, effective, and safer rehabilitation exercises.

With that, let's proceed to the next question. What else would you like to learn about chatbots and their role in sports rehabilitation?

G

How are chatbots improving the psychological health of injured athletes? Can they really pick up on the emotional tone in our words and adjust their responses to suit our mood?

Henry: A fascinating question indeed, one that delves into the ability of chatbots to understand and respond to human emotions. Let's start with Dr. Perry, our sports psychologist.

Dr. Perry: Thank you, Henry. In the psychological aspect of sports rehabilitation, chatbots can indeed play a crucial role. They can provide motivational messages, remind athletes of their progress, and offer cognitive-behavioral strategies to manage pain and anxiety. As for picking up on the emotional tone in words, chatbots, powered by Natural Language Processing and sentiment analysis algorithms, can indeed identify certain emotions from text input. However, it's important to note that this technology is still evolving and, while promising, it may not always accurately reflect the complexity and nuances of human emotion.

Henry: Intriguing insights, Dr. Perry. Nelson, could you share some light on the technical aspects of this question?

Nelson: Sure, Henry. Dr. Perry hit the nail on the head. Sentiment analysis is a field within Natural Language Processing that deals with identifying and extracting subjective information from source materials. Advanced AI models are getting better at picking up cues from the language we use and can often accurately determine the sentiment behind the text. That being said, it's a challenging task as human emotions are complex and nuanced. There's ongoing research in the field and the technology is consistently improving. So, while chatbots can already adjust their responses to a certain degree, we can expect them to get even better at this in the future.

Henry: Thanks, Dr. Perry and Nelson. To summarize, chatbots can support the psychological health of injured athletes in several ways, including motivation, progress reminders, and anxiety management. They can, to some extent, detect the emotional tone of an athlete's words using sentiment analysis, a subset of Natural Language Processing, and adjust their responses accordingly. However, this technology is still evolving and, while it shows promise, it doesn't perfectly capture the full range of human emotions. Let's move on to the next question.

G

How can chatbots improve in nutrition management for athletes during recovery?

Henry: That's a fantastic question which brings us to the intersection of nutrition and technology in sports rehabilitation. Nikki, as our sports nutritionist, could you provide some insights on this?

Nikki: Absolutely, Henry. Nutrition is a key element of an athlete's recovery. Chatbots can already provide basic dietary advice based on the athlete's current stage of recovery and activity level. They can suggest meal plans, remind athletes to stay hydrated, and even track their daily nutrient intake. However, there's still a lot of room for improvement. Ideally, chatbots could be integrated with wearable devices or health tracking apps to monitor the athlete's energy expenditure and make real-time dietary suggestions. Moreover, advances in AI could make chatbots more adaptable, allowing them to cater to various dietary preferences, intolerances, or allergies, resulting in a truly personalized nutritional plan.

Henry: Thanks, Nikki. Nelson, can you add something from a technical standpoint?

Nelson: Nikki is spot on. Integrating chatbots with other digital tools like wearable devices or health tracking apps can make the process of nutrition management much more accurate and personalized. It's all about the data - the more quality data the chatbot has access to, the better advice it can provide. And with the advancement in machine learning algorithms, the chatbots can learn from each interaction, continually improving their dietary suggestions.

Henry: Thanks, Nikki and Nelson. To summarize, chatbots can already aid significantly in nutrition management during an athlete's recovery. They can provide dietary advice, suggest meal plans, and remind athletes to hydrate. By integrating chatbots with other tools like wearable devices and health apps, we can achieve even more personalized nutrition management. As AI and machine learning technologies advance, chatbots will continue to improve, offering more accurate and personalized nutritional advice. Now, let's move to the next question.

G

How do chatbots keep track of an athlete's recovery? And do they have any special advantages over the old ways we used to do this?

Henry: That's an excellent question. It's about the capabilities of chatbots to track an athlete's recovery and how they might offer advantages over traditional methods. Dr. Mandy, as a sports physiotherapist, could you share your insights?

Dr. Mandy: Of course, Henry. Chatbots can keep track of an athlete's recovery by recording and analyzing data that the athlete provides during their interactions. This could include details about their pain levels, exercise compliance, sleep quality, and more. The chatbot can then use this data to monitor progress, adjust exercise or recovery plans, and provide feedback. Compared to traditional methods, this could offer several advantages. It allows for continuous, real-time tracking, which can lead to more timely adjustments in the recovery plan. It also helps in maintaining a comprehensive and accurate record of the athlete's recovery journey.

Henry: Thanks, Dr. Mandy. Nelson, could you add to this from a technological perspective?

Nelson: Sure, Henry. Dr. Mandy covered it well. From a technology standpoint, chatbots leverage machine learning algorithms to analyze data and identify patterns or trends, which can provide insights into an athlete's recovery. Moreover, since chatbots are typically cloud-based, the data is stored securely and can be accessed anytime, from anywhere. This not only provides flexibility but also ensures that the athlete's progress is tracked even if they move or change their healthcare provider.

Henry: Thank you both for your insights. In summary, chatbots can keep track of an athlete's recovery by consistently recording and analyzing data provided by the athlete. This allows for real-time tracking and the ability to adjust recovery plans promptly. Compared to traditional methods, chatbots offer the advantages of continuous monitoring, accurate record-keeping, and flexibility of access. Let's move on to our next question.

G

When we bring chatbots into athlete recovery, how do we make sure we respect patient choice, keep data private and safe? How do we handle worries about things like bias, fairness, and not undervaluing the human touch in our clinics?

Henry: Those are crucial points to consider when implementing chatbots in the realm of sports rehabilitation. Ensuring patient choice, data privacy, fairness, and maintaining the irreplaceable human touch are all important aspects. Let's discuss this, starting with Nelson, our expert in Natural Language Processing for clinical application.

Nelson: Thank you, Henry. The issues of patient choice, data privacy, and bias are indeed critical. Firstly, patient choice must always be respected. Chatbots should be offered as a supplement to traditional care, not a replacement. Secondly, regarding data privacy, chatbots should be designed and deployed following strict data privacy regulations like GDPR or HIPAA, depending on the region. The data must be encrypted, and only authorized personnel should have access to it.

In terms of bias and fairness, this is a challenge in AI that we're continuously working on. Training data should be diverse and representative to avoid biases in the chatbot's responses. And there should be transparency about how the AI makes decisions, known as 'explainable AI'.

Henry: Thanks, Nelson. Dr. Mandy, how about the concern of undervaluing the human touch in our clinics?

Dr. Mandy: That's a fundamental concern, Henry. Technology, no matter how advanced, can't replace the human touch, empathy, and judgement that healthcare professionals bring. Chatbots should be used as tools that complement and enhance our work, not replace us. We should ensure that our patients always have access to human interaction and support whenever they need it.

Henry: Thank you, Nelson and Dr. Mandy. In summary, ensuring patient choice, data privacy, fairness, and maintaining the human touch are all critical when implementing chatbots in sports rehabilitation. Chatbots should be introduced as a supplementary tool, not a replacement for human care. Strict data privacy regulations must be adhered to, and efforts must be made to eliminate bias from AI systems. Despite the advancement of technology, the irreplaceable human touch, empathy, and judgement that healthcare professionals bring should always be valued. Let's move on to our next question.

G

because the time is up, please think of one last question to ask the panel to conclude the discussion.

Henry: Thank you, audience, for your thoughtful questions. As we're nearing the end of our time together, let's conclude with one final question for our panel. Considering all that we've discussed today, what do you each believe is the most exciting opportunity or development for the use of chatbots in sports rehabilitation in the near future? We'll start with Dr. Mandy and go down the line.

G

o Experts on the panel, please provide comments on the question.

Dr. Mandy: The future of chatbots in sports rehabilitation is undoubtedly exciting. I'm particularly enthusiastic about the potential for integrating chatbots with sensor technology and real-time monitoring tools. It could allow us to provide instant feedback to athletes about their movements and techniques, making rehabilitation more effective and reducing the risk of re-injury.

Dr. Perry: For me, it's the potential for chatbots to provide psychological support to athletes. As the technology improves, chatbots could become more adept at providing tailored mental health interventions, from stress management techniques to motivational messages. This could greatly enhance the holistic support we provide to athletes during their recovery.

Nikki: As a nutritionist, I'm excited about the potential of chatbots for more personalized and dynamic nutrition management. Integrated with health tracking apps or wearable devices, chatbots could provide real-time dietary advice based on an athlete's current energy expenditure and recovery stage.

Allan: From an athlete's perspective, I look forward to the convenience and constant support that chatbots can provide. They can be available 24/7, providing answers to our questions and offering support when human therapists are not immediately available. This could greatly improve the recovery experience for many athletes.

Nelson: Technologically speaking, the ongoing advancements in Natural Language Processing and machine learning are incredibly exciting. These advancements will not only make chatbots more intelligent and responsive but also help them better understand and respond to the nuanced language and emotions of the users, leading to a more personalized and empathetic user experience.

Henry: Thank you all for sharing your insights. It's clear that the future of chatbots in sports rehabilitation holds many promising developments, from real-time monitoring and feedback to tailored mental health interventions, dynamic nutrition management, and more personalized user experiences. As we continue to advance and refine these technologies, we're opening up new possibilities for supporting athletes on their recovery journeys. Thank you to our esteemed panel and our audience for this engaging and insightful discussion.
