## Supplementary File 4 for "Interdisciplinary Inquiry via PanelGPT: Application to Explore Chatbot Application in Sports Rehabilitation"

G

How are chatbots improving the psychological health of injured athletes? Can they really pick up on the emotional tone in our words and adjust their responses to suit our mood?

Yes, chatbots are playing an increasingly important role in various sectors, including mental health. For injured athletes, chatbots can offer a range of benefits, especially in terms of psychological support. Let's consider some of the ways in which chatbots are contributing to this:

1. **Constant Availability**: Chatbots are available 24/7, meaning they can offer support and comfort to athletes at any time of day or night. This is particularly valuable for individuals who may be dealing with feelings of isolation or anxiety in the early hours of the morning when human support may not be readily available.
2. **Emotion Recognition**: Some sophisticated chatbots, like advanced versions of GPT (like GPT-4 and beyond), use machine learning to understand the emotional tone in a user's words. They can then adjust their responses to suit the user's mood. This involves natural language processing, sentiment analysis, and advanced algorithms that can "read" and understand language on a deeper level. It's not perfect and doesn't have the full emotional understanding and nuance of a human, but it has been shown to be helpful.
3. **Guided Meditation and Relaxation**: Chatbots can lead users through guided meditation and relaxation exercises, which can be particularly beneficial for athletes recovering from injuries. They can aid in stress management and help individuals maintain a positive outlook.
4. **Goal Setting and Progress Tracking**: Chatbots can help athletes set and track recovery goals, offering encouragement and positive reinforcement along the way. This can contribute to a sense of progress and forward momentum, which can be particularly beneficial in terms of maintaining mental health during a lengthy recovery period.
5. **Providing Reliable Information**: Chatbots can also provide reliable, evidence-based information about recovery from injuries. This can help to reduce anxiety and promote a sense of control.
6. **Reducing Stigma**: Many people, athletes included, might feel a stigma around discussing mental health. A chatbot can offer a judgment-free zone to express feelings and concerns.
7. **Therapeutic Support**: Cognitive behavioral therapy (CBT) chatbots have been developed and have shown effectiveness in delivering mental health support. They guide users through cognitive-behavioral techniques to manage stress, anxiety, and other negative feelings.

However, it is important to remember that while chatbots can offer a useful tool in supporting mental health, they should not replace professional mental health services. Athletes struggling with their mental health should reach out to qualified mental health professionals for support. It's also crucial that such tools are designed and used ethically, with appropriate measures in place to protect users' privacy and data.

G

How are chatbots improving the psychological health of injured athletes? Can they really pick up on the emotional tone in our words and adjust their responses to suit our mood?

Yes, chatbots can play an instrumental role in improving the psychological health of injured athletes, and they're becoming increasingly sophisticated at understanding and responding to emotional tones in the text they process. Here are a few ways that chatbots are currently being used to support the mental health of injured athletes:

1. **Emotional Support**: Chatbots can be programmed to provide emotional support by acknowledging an athlete's feelings, validating their experience, and suggesting helpful coping mechanisms. The chatbot can ask about an athlete's current mood and feelings, and offer empathetic responses.
2. **Cognitive Behavioral Therapy (CBT) Techniques**: Some chatbots incorporate principles from CBT to help athletes manage stress and anxiety. They can teach athletes to recognize negative thought patterns, challenge these thoughts, and replace them with healthier ones.
3. **Information and Resources**: Chatbots can provide information about the injury, recovery process, and available resources. This can reduce uncertainty and anxiety, helping the athlete feel more in control of their situation.
4. **Guided Meditations and Breathing Exercises**: Some chatbots guide users through mindfulness exercises, which can help reduce stress and improve mood.
5. **Encouragement and Motivation**: Chatbots can provide regular encouragement and motivational messages, helping athletes maintain a positive mindset throughout their recovery.

Regarding your question about chatbots detecting emotional tone, advancements in natural language processing (NLP) and machine learning have made it possible for chatbots to analyze the emotional content of a text. This technology, known as sentiment analysis or emotion AI, can identify and understand nuances in language, such as tone, context, and sentiment. This enables the chatbot to adjust its responses based on the perceived emotional state of the user. While this technology has made significant strides, it's important to note that it's not perfect and sometimes may not fully grasp the complexity and subtlety of human emotions.

Overall, while chatbots can't replace human therapists or psychologists, they can provide valuable support and resources, particularly in situations where an athlete may not have immediate access to professional mental health services.

G

How are chatbots improving the psychological health of injured athletes? Can they really pick up on the emotional tone in our words and adjust their responses to suit our mood?

Chatbots, including those driven by AI like me, are playing an increasingly important role in supporting mental health, including the psychological health of injured athletes. Here are a few ways how they are helping:

1. **Availability and Accessibility:** AI-powered chatbots are available 24/7, allowing athletes to access psychological support whenever they need it, even if it's in the middle of the night or during times when a human therapist may not be available.
2. **Anonymity:** Some people, including athletes, might feel stigmatized by seeking mental health support. Chatbots provide a level of anonymity that can make people feel more comfortable discussing their feelings.
3. **Consistency:** Chatbots provide consistent responses which can be helpful in managing mental health. They don't suffer from mood swings, fatigue, or other human factors that might influence the quality of support.
4. **Techniques and Exercises:** Chatbots can provide psychological techniques and exercises, such as Cognitive Behavioral Therapy (CBT) techniques, mindfulness exercises, and more to help athletes manage their mental health.
5. **Monitoring Progress:** Chatbots can help monitor an athlete's mental health progress over time, and provide insights and analysis that can be useful for both the athlete and their support team.

As for picking up on the emotional tone of users' inputs, yes, advanced AI chatbots are increasingly able to do this through a process called sentiment analysis or emotional AI (also known as affective computing). This involves using machine learning to analyze the emotional tone of a user's inputs and adjust responses accordingly. However, it's important to note that while these systems are improving, they are not perfect and may not always accurately interpret the emotional tone of inputs. AI doesn't truly understand emotions in the way humans do, but instead, they identify patterns that correspond to different emotional states.

Furthermore, it's important to note that while chatbots can provide valuable support, they are not a replacement for professional mental health services. Athletes dealing with significant psychological challenges should seek help from a qualified professional. Chatbots are an additional tool that can complement but not replace traditional mental health care.
