## Supplementary File 5 for "Interdisciplinary Inquiry via PanelGPT: Application to Explore Chatbot Application in Sports Rehabilitation"

Model: GPT-4

G

How can chatbots help in an athlete's recovery? Can they really understand things like how the athlete moves, or how they distribute their weight, and then give specific advice about exercises that are just right for them?

As of my knowledge cutoff in September 2021, AI chatbots, like ChatGPT, can't directly understand or observe physical activity, such as how an athlete moves or distributes their weight. They don't have vision or physical senses. Their abilities are largely limited to text-based interactions.

However, AI systems could be a helpful part of an athlete's recovery process in a few ways:

1. **Injury Information and Guidance:** AI chatbots can provide general information about various injuries, their symptoms, and common recovery processes. This might help athletes understand what they're experiencing, although the information should not be taken as medical advice.
2. **Exercise and Rehabilitation Information:** AI chatbots can share common exercises or rehabilitation activities recommended for certain injuries. However, since a chatbot can't observe an athlete's form or provide specific feedback, these should be used in conjunction with advice from a physical therapist or other health professional.
3. **Reminders and Tracking:** AI systems can help athletes remember when to do exercises, track their progress over time, and remind them about appointments.
4. **Motivation and Mental Health Support:** Many athletes find the recovery process frustrating or mentally challenging. AI chatbots can provide motivational messages, guided relaxation exercises, and other forms of emotional support.

As AI and technology continue to advance, the ways in which they can assist athletes will likely expand. For example, future integration with wearable technology that can track movement, heart rate, and other physiological parameters could potentially provide a more personalized advice and rehabilitation process. Such systems might use AI to analyze data from these devices and provide feedback or suggestions, but that technology is still developing and would need careful design and oversight from health professionals to ensure it is accurate and safe.

In all cases, an AI should be considered as an adjunct tool and should not replace advice from healthcare professionals who can provide personalized and accurate advice based on direct observations and interactions.

Model: GPT-4

G

How can chatbots help in an athlete's recovery? Can they really understand things like how the athlete moves, or how they distribute their weight, and then give specific advice about exercises that are just right for them?

Chatbots, as of my knowledge cutoff in September 2021, aren't able to directly observe or measure physical characteristics or movements such as how an athlete moves or how they distribute their weight. However, chatbots can indeed play a significant role in an athlete's recovery and conditioning process in the following ways:

1. **Information Provider**: Chatbots can provide detailed information about various types of exercises, their benefits, common mistakes, and how to perform them correctly. This can be particularly useful if an athlete is learning new exercises or training routines.
2. **Exercise Schedule Management**: Chatbots can help manage exercise schedules and remind athletes when it's time to exercise, rest, hydrate, and eat. This can be crucial for maintaining an optimal recovery and training regimen.
3. **Injury Advice**: If an athlete has been injured, a chatbot can provide basic advice on how to manage the injury. However, this should not replace professional medical advice.
4. **Data Collection**: If the athlete uses any wearable devices or tracking apps, the chatbot can utilize the collected data (such as heart rate, step count, etc.) to offer personalized advice.
5. **Psychological Support**: Athletes undergoing recovery often experience psychological stress. Chatbots can provide emotional support and motivation, helping them to maintain a positive mindset.

In the future, there may be more sophisticated technologies and AI models that can analyze video to understand an athlete's movements and provide personalized advice based on those observations. This is an active area of research and development, so we may see these capabilities in the future.

Remember that while chatbots can provide valuable support, they are not a replacement for professional advice from coaches, physical therapists, nutritionists, and doctors. Always consult with a healthcare professional for medical advice.

Model: GPT-4

G

How can chatbots help in an athlete's recovery? Can they really understand things like how the athlete moves, or how they distribute their weight, and then give specific advice about exercises that are just right for them?

Chatbots can certainly be of great help in an athlete's recovery, especially in the aspects of monitoring, motivation, reminders, providing general advice and much more. However, it's important to remember that as of my last update in September 2021, chatbots like me, even the advanced AI models, don't have the ability to visually observe physical movement or weight distribution directly, nor can we physically assess an individual's condition. We depend on the data provided to us.

Let me expand a bit more on the ways chatbots can assist in an athlete's recovery:

1. **Monitoring progress**: Chatbots can collect and analyze data over time, based on the user's input. For example, athletes can report their daily pain levels, range of motion, or other relevant metrics. The chatbot can then track this data to help monitor the athlete's progress.
2. **Motivation and adherence**: Chatbots can remind athletes to perform their exercises or take their medication at appropriate times, enhancing adherence to rehabilitation plans. They can also provide motivational messages and goal-setting features to help athletes stay committed to their recovery process.
3. **Educational resources**: Chatbots can provide athletes with a wealth of information related to their injury and recovery. This could include explanations of their injury, why certain exercises are beneficial, dietary recommendations, and more.
4. **Telemedicine applications**: In situations where a telemedicine integration is possible, chatbots could serve as an intermediary between athletes and their healthcare providers. This could facilitate remote consultations, early detection of complications, and so on.

For more sophisticated understanding and advice such as assessing how an athlete moves, how they distribute their weight, or giving highly specific advice about exercises, current technologies as of 2021 are leveraging a combination of AI, IoT (Internet of Things) devices, and sensors. For example, wearable devices or smart gym equipment could potentially collect data on an athlete's movements, biomechanics, and weight distribution. These data could then be analyzed using AI techniques to provide specific feedback and advice.

Such systems would likely involve collaboration between AI models, healthcare professionals, and sports scientists to ensure the advice is accurate, effective, and safe. The AI could help identify patterns and trends in the data, while the human experts could interpret these findings and use their professional judgment to provide the best advice.

Please note that such solutions need careful design, including strict privacy measures and should be used under professional supervision as incorrect advice could lead to injuries or slow down the recovery process. And lastly, advancements in this area are fast and new functionalities might have been developed since my last update. Always refer to the most recent sources for the latest information.
