## Supplementary File 6 for "Interdisciplinary Inquiry via PanelGPT: Application to Explore Chatbot Application in Sports Rehabilitation"

JC

You will assume various roles in a thought-provoking panel on " Chatbots in Sports Rehabilitation: Opportunities and Challenges." The simulation will involve a skilled moderator, Henry, who will guide the discussion, along with esteemed professionals in their respective fields: Dr. Mandy, a sports physiotherapist; Dr. Perry, a sports psychologist; Nikki a sports nutritionist; Allan, a professional athlete who had successfully navigated the rehabilitation process after a severe injury; and Nelson, an expert in Natural Language Processing for clinical application. I will role-play the audience and send you questions or reminders. While multiple experts can offer their insights, no question should be left unanswered. A BING search may be performed in order to answer each of the questions with the most up to date information. When replying to a question, please use spoken English. Henry, please begin the panel discussion by introducing the panel, followed by asking the panel about the role of chatbots in sports rehabilitation and how they see this role evolving.

(Henry): Good afternoon, everyone. I'd like to extend a warm welcome to our esteemed panelists and our audience. We are gathered here today to engage in a thought-provoking conversation on "Chatbots in Sports Rehabilitation: Opportunities and Challenges."

Firstly, allow me to introduce our panel. We have Dr. Mandy, a highly respected sports physiotherapist known for her innovative techniques in aiding athletes' recovery. Also, we have Dr. Perry, a sports psychologist whose work has been instrumental in understanding the mental fortitude required in sports. Then, we have Nikki, a sports nutritionist whose expertise in diet and recovery has been acknowledged globally. Allan, a professional athlete, is here to provide insights from his personal journey of recovery from a severe injury. Lastly, we have Nelson, an expert in Natural Language Processing for clinical applications, who brings a unique perspective to our discussion.

Now, moving on to the main topic for today. Let's delve into the role of chatbots in sports rehabilitation. As we know, the advent of artificial intelligence and machine learning has brought about revolutionary changes across various sectors, and sports rehabilitation is no exception.

To kick off the conversation, I'd like to ask our esteemed panel: What role do you see chatbots playing in sports rehabilitation currently, and how do you envision this role evolving in the future?

JC

Experts on the panel, please provide comments on the questions.

(Dr. Mandy): Thank you, Henry. From my perspective as a sports physiotherapist, chatbots have been proving extremely useful. They can provide immediate responses to athletes' inquiries about their rehabilitation exercises, ensure they're performing them correctly, and even monitor their progress over time. This offers a level of interaction and feedback that would be impossible for me to provide constantly.

However, as we look towards the future, I think the real potential lies in chatbots' ability to provide personalized rehabilitation programs. By analyzing an athlete's injury, progress, and response to different exercises, chatbots could potentially design and modify rehabilitation plans in real time. It's truly an exciting prospect.

(Dr. Perry): I echo Dr. Mandy's sentiments. As a sports psychologist, I see immense potential for chatbots in providing mental health support. Rehabilitation can be a stressful and isolating process. Having a chatbot available 24/7 for conversation and support can help athletes feel less alone. In the future, I see chatbots equipped with advanced sentiment analysis capabilities to provide even more nuanced emotional support.

(Nikki): From a nutritional standpoint, chatbots can provide immediate and personalized dietary advice to athletes. They can help track caloric intake, suggest meal plans, and even remind athletes to hydrate or eat at regular intervals. As technology advances, I envision chatbots integrated with wearable tech, adjusting dietary advice based on the athlete's real-time biometrics.

(Allan): As someone who has gone through the rehabilitation process, the prospect of having a chatbot companion is exciting. When I was rehabilitating, I often had questions or concerns outside of my scheduled appointments. A chatbot can provide immediate responses and reassurances. In the future, I think chatbots can also play a crucial role in motivating and inspiring athletes during their recovery journey.

(Nelson): From a technology perspective, chatbots are continually evolving. Today, they can handle basic tasks such as answering questions or tracking progress. But with advancements in natural language processing and machine learning, chatbots will become even more sophisticated. They'll be able to understand context, infer meaning from incomplete information, and even predict user needs before they're expressed. This opens up incredible possibilities for sports rehabilitation, from providing more personalized care to detecting early signs of potential issues in an athlete's rehabilitation process.

JC

Henry, would you summarize and open the floor for questions.

(Henry): Absolutely, and thank you all for your insightful comments. It's clear that chatbots have already begun to play a significant role in sports rehabilitation, providing immediate responses, monitoring progress, providing emotional support, and offering personalized dietary advice. As we move towards the future, the potential of chatbots in this field is vast. With advancements in technology, they could provide even more personalized care, motivate athletes, and anticipate needs before they're expressed.

It's truly an exciting time, and I am eager to see how these developments unfold. At this point, I'd like to open the floor for questions. Audience members, please feel free to ask your questions to our panel.

JC

Audience: What education should we offer to guide our athletes in the use of a chatbot for rehab?

Finished browsing

To guide athletes in using chatbots for rehab, we must consider a holistic approach to education that goes beyond just teaching them how to use the tool. Here are some key areas to consider:

**Understanding Chatbots:** It's essential to explain what a chatbot is and how it works. A chatbot is a computer program that engages the user in conversation, either through text or speech. It responds to set commands, and the more advanced ones use machine learning, learning from each interaction, detecting preferences, and making recommendations based on past requests[1]. This understanding will help athletes trust and interact effectively with the chatbot.

**Embed Chatbots in Familiar Platforms:** To ease the learning curve, we can embed a chatbot within an existing app that athletes already use, acting as the first line of response to incoming requests[1].

**Integrating Chatbots into Learning Management Systems (LMS):** Chatbots can connect learners to your organization's Learning Management System (LMS) not simply to request learning, but to select and recommend a particular piece of training and to build a learner profile based on learner interaction and perceived need[1].

**Real 'Just-In-Time' Learning:** The application of AI to performance support (PS) can deliver training at the point of need, making it more relevant and cost-effective. This approach will allow athletes to benefit from improved performance and efficiency with real 'just-in-time' learning[1].

**Organizational Readiness:** Ensure your organization is ready to take full advantage of this new enhanced Performance Support. It's not just about installing new technology or reconfiguring your existing systems, but also about learning design, organizational structure, and workflow processes[1].

**Performance Support Maturity:** Consider the extent to which performance support is embedded in the workflow, the role of your IT department, how PS might start in a specific area but needs to proliferate into other areas, the integration of performance support materials with training, and the relationship between training and performance support[1].

**Performance Support Needs:** Make sure the information and support provided are specific to athletes' needs and outcomes. Information and solutions need to be chunked and packaged with the watchwords being 'just enough' and 'just in time'. Measure its effect on performance and refine it based on experience and need[1].

Lastly, perhaps the biggest change you need to make is how you approach training and learning in your organization[1]. Dr. Mandy, could you add more details from the perspective of sports physiotherapy?
