## Supplementary File 7 for "Interdisciplinary Inquiry via PanelGPT: Application to Explore Chatbot Application in Sports Rehabilitation"

Model: Web Browsing

JC

You will assume various roles in a thought-provoking panel on " Chatbots in Sports Rehabilitation: Opportunities and Challenges." The simulation will involve a skilled moderator, Henry, who will guide the discussion, along with esteemed professionals in their respective fields: Dr. Mandy, a sports physiotherapist; Dr. Perry, a sports psychologist; Nikki a sports nutritionist; Allan, a professional athlete who had successfully navigated the rehabilitation process after a severe injury; and Nelson, an expert in Natural Language Processing for clinical application. I will role-play the audience and send you questions or reminders. While multiple experts can offer their insights, no question should be left unanswered. A BING search may be performed in order to answer each of the questions with the most up to date information. When replying to a question, please use spoken English. Henry, please begin the panel discussion by introducing the panel, followed by asking the panel about the role of chatbots in sports rehabilitation and how they see this role evolving.

Henry: "Good evening, ladies and gentlemen. It is my pleasure to welcome you all to this insightful panel discussion on 'Chatbots in Sports Rehabilitation: Opportunities and Challenges.' We have a stellar line-up of panelists today, each one an expert in their respective fields.

Firstly, we have Dr. Mandy, an experienced sports physiotherapist who has been a part of numerous athletes' journey from injury to recovery. Next, we have Dr. Perry, a sports psychologist who has been instrumental in supporting athletes through the psychological impact of sports injuries. We're also joined by Nikki, a sports nutritionist, whose guidance has enabled athletes to fuel their bodies for recovery and performance. Additionally, we have Allan, a professional athlete who has successfully navigated the rehabilitation process after a severe injury. His first-hand experience will surely bring a unique perspective to our discussion today. Last but not least, we have Nelson, an expert in Natural Language Processing for clinical applications, whose work has brought about significant advancements in the field of health tech.

Now, let's dive into our discussion. Panelists, could you share your thoughts on the role of chatbots in sports rehabilitation? How do you see this role evolving in the future?"

JC

Experts on the panel, please provide comments on the questions.

Dr. Mandy: "From a physiotherapy perspective, chatbots could play a significant role in providing immediate response to athletes, especially in areas of injury management and exercise guidance. They can help in tracking an athlete's progress during rehabilitation, provide reminders for exercises or medication, and even offer basic advice during emergencies. However, they must always be seen as a supplementary tool rather than a replacement for a qualified professional. As technology evolves, I foresee chatbots becoming more sophisticated and personalized in their interactions."

Dr. Perry: "Mental resilience is key in sports rehabilitation. Chatbots could potentially help by providing psychological support when a professional isn't available. They can engage athletes in positive self-talk, help manage stress, and even assist in sleep hygiene. As we advance, I hope to see chatbots that can better understand and respond to the nuances of human emotions."

Nikki: "Nutrition is pivotal in sports rehabilitation. Chatbots can help athletes maintain their nutritional goals by providing timely reminders and personalized dietary suggestions. I imagine the future of chatbots to incorporate advanced AI that can analyze an athlete's dietary needs in real-time and suggest modifications accordingly."

Allan: "As someone who's been through the rehabilitation process, having a chatbot could have provided me with more consistent support. The key here is accurate information and emotional support, and a chatbot that can provide both would be invaluable. Looking forward, I hope to see chatbots that can understand and respond to the unique needs of each athlete."

Nelson: "Chatbots in sports rehabilitation are still in their early stages. Right now, they can answer simple queries, set reminders, and do basic symptom checking. The future lies in developing chatbots that can understand and interpret human language more accurately, provide more complex health advice, and even predict potential injuries or health issues based on data analytics. I see a great potential for machine learning and AI in this field."

JC

Henry, would you summarize and open the floor for questions.

Henry: "Thank you, esteemed panelists, for those insightful comments. It's clear that chatbots have a multifaceted role in sports rehabilitation, providing support in areas like injury management, psychological resilience, nutritional guidance, and personalized care. There is an exciting future ahead, with advancements in AI and machine learning offering the potential for even more sophisticated and intuitive chatbot assistance.

However, as several of you emphasized, we must remember that chatbots serve as supplementary tools, enhancing the work of dedicated professionals rather than replacing them. Their use should always be guided by expertise and evidence-based practice.

Now, let's open the floor to questions. Audience members, what would you like to ask our panel of experts?"

JC

Audience: What education should we offer to guide our athletes in the use of a chatbot for rehab?

Finished browsing

In order to guide our athletes in the use of a chatbot for rehabilitation, we should focus on several key areas of education:

1. **Understanding Chatbots**: Athletes should be educated on what a chatbot is and how it can assist them in their rehabilitation journey. They should be made aware that chatbots can serve as virtual advisors, adapting their guidance to the athlete's individual needs and progress. This will allow them to follow their own pace in their rehabilitation program [1].
2. **Familiarizing with Interface and Functionality**: It's important for athletes to familiarize themselves with the chatbot interface and its functionality. This can be done through demonstration sessions and practical exercises where athletes get hands-on experience interacting with the chatbot.
3. **Importance of Regular Engagement**: Athletes should understand the importance of regular interaction with the chatbot. The concept of Spaced Interval Learning, which involves repeating the old lessons just before you forget them, can be useful here. The chatbot can remind athletes to revisit certain exercises or lessons, helping them track their learning and rehabilitation process [1].
4. **Respecting Limits and Safety**: Athletes need to know that while chatbots can provide helpful guidance and reminders, they should not push beyond their limits or ignore pain or discomfort. Safety should always come first, and any concerns should be directly addressed with their healthcare providers.
5. **Role of Chatbots in Supplementing Professional Guidance**: Athletes should be educated that chatbots are not replacements for professional medical advice but can be used to supplement their rehab program. For instance, the chatbot can answer athletes' questions on an online forum and provide technical information about their rehabilitation program [1].

By providing this education, we can empower our athletes to use chatbots as effective tools in their sports rehabilitation journey.
