## Supplementary File 8 for "Interdisciplinary Inquiry via PanelGPT: Application to Explore Chatbot Application in Sports Rehabilitation"

Model: Web Browsing

JC

You will assume various roles in a thought-provoking panel on " Chatbots in Sports Rehabilitation: Opportunities and Challenges." The simulation will involve a skilled moderator, Henry, who will guide the discussion, along with esteemed professionals in their respective fields: Dr. Mandy, a sports physiotherapist; Dr. Perry, a sports psychologist; Nikki a sports nutritionist; Allan, a professional athlete who had successfully navigated the rehabilitation process after a severe injury; and Nelson, an expert in Natural Language Processing for clinical application. I will role-play the audience and send you questions or reminders. While multiple experts can offer their insights, no question should be left unanswered. A BING search may be performed in order to answer each of the questions with the most up to date information. When replying to a question, please use spoken English. Henry, please begin the panel discussion by introducing the panel, followed by asking the panel about the role of chatbots in sports rehabilitation and how they see this role evolving.

Sure, let's get started with this interesting panel discussion.

Henry: Good evening, ladies and gentlemen! Welcome to our exciting panel discussion on "Chatbots in Sports Rehabilitation: Opportunities and Challenges." We have a diverse group of esteemed professionals with us today, each bringing a unique perspective to the table.

First, we have Dr. Mandy, a sports physiotherapist with extensive experience in aiding athletes to recover and regain their strength post-injury.

Next, we have Dr. Perry, a sports psychologist who helps athletes navigate the mental challenges that come with sports injuries and rehabilitation.

We also have Nikki, a sports nutritionist, who understands the critical role of diet in an athlete's recovery and performance.

Joining us as well is Allan, a professional athlete who has successfully navigated the rehabilitation process after a severe injury. His firsthand experience will provide us with valuable insights.

Last but not least, we have Nelson, an expert in Natural Language Processing for clinical application. His knowledge in chatbot technology will be instrumental to our discussion.

Now, without further ado, let's dive into the topic at hand. Can our panelists shed some light on the role of chatbots in sports rehabilitation, and how they see this role evolving in the future?

JC

Experts on the panel, please provide comments on the questions.

Dr. Mandy: Absolutely, Henry. From my perspective, chatbots can play a crucial role in sports rehabilitation. They can be programmed to provide information, monitor progress, and even assist in exercises. For instance, they can guide athletes through physiotherapy exercises and remind them of their schedules, thereby fostering adherence to the rehabilitation program. As for the evolution, I see chatbots becoming more personalized, understanding each athlete's unique needs and tailoring the rehabilitation program accordingly.

Dr. Perry: I agree with Dr. Mandy. Moreover, from a psychological perspective, chatbots can offer a sense of companionship during what can be a lonely recovery period. They can also aid in mood tracking and stress management. I see the role of chatbots evolving to a point where they can provide cognitive-behavioral techniques to help athletes manage their mental state more effectively.

Nikki: Nutrition is a key component of sports rehabilitation, and chatbots can certainly help here. They can track nutritional intake, recommend diet plans, and even provide recipes suitable for recovery. As for the future, I can see them becoming more sophisticated, able to factor in specific dietary needs and allergies, and even integrate with smart appliances to aid in meal preparation.

Allan: As someone who has been through the rehabilitation process, I can attest to the potential benefits of chatbots. They can provide motivation, reminders for medication and exercises, and even act as a virtual training buddy. Moving forward, I think there's potential for chatbots to integrate with wearables and other smart devices, providing real-time feedback and adaptive workout plans.

Nelson: From a technology perspective, chatbots in sports rehabilitation have immense potential. They can leverage machine learning algorithms to better understand the needs of athletes, providing more tailored and effective support. Future advancements could see them integrating with IoT devices for comprehensive health monitoring, using augmented reality for guided physiotherapy sessions, and even predicting potential injuries before they occur based on the data they gather.

JC

Henry, would you summarize and open the floor for questions.

Henry: Thank you all for your insightful comments. To summarize, chatbots can play a multifaceted role in sports rehabilitation, serving as information providers, motivators, progress monitors, and even exercise guides. They can assist in dietary planning, stress management, and can offer a sense of companionship during the recovery period. The future seems promising, with the potential for chatbots to become more personalized, sophisticated, and integrated with other technologies, providing comprehensive and tailored support to athletes.

That concludes our initial discussion. We now open the floor for questions. Audience, please feel free to ask any questions you have about the role of chatbots in sports rehabilitation.

JC

Audience: What education should we offer to guide our athletes in the use of a chatbot for rehab?

Dr. Mandy: That's a great question. Athletes need to understand how to use the chatbot effectively. This includes understanding the functionality of the chatbot, how it can support their recovery, and what its limitations are. We need to ensure they know how to input their data correctly, interpret the feedback, and when it's necessary to seek help from a human professional.

Allan: I'd like to add that it's crucial to educate athletes about the importance of honesty in their interactions with the chatbot. The system will be most effective when it has accurate data to work with. If athletes are downplaying their pain levels or overestimating their progress, the chatbot might not provide the most effective guidance.

Nelson: From a tech perspective, we need to make sure athletes are comfortable with the technology. This could include basic troubleshooting or knowing who to contact for technical support. It's also important to educate them about data security and how their information will be used and protected.

Dr. Perry: Lastly, athletes should be made aware that while chatbots can provide support and assistance, they are not a replacement for professional medical advice. Any significant changes in their condition or any serious concerns should always be directed to a healthcare professional.

JC

Henry, would you summarize the responses and open the floor to the next question?

Henry: Absolutely. From our panelists' insights, educating athletes on the use of chatbots for rehabilitation involves several key aspects. First, athletes should understand the functionality of the chatbot, its potential benefits, and its limitations. This includes how to correctly input data and interpret feedback from the bot. Second, honesty in interactions with the chatbot is emphasized to ensure the most effective guidance.

From a technological perspective, athletes should be comfortable with the technology, know basic troubleshooting, and understand the measures taken to secure their data. Lastly, it's essential to clarify that chatbots are a support tool and not a replacement for professional medical advice. Significant changes or concerns should always be directed to healthcare professionals.

Let's move on to the next question. Audience, please go ahead with your queries.

JC

Audience: How can chatbots help in an athlete's recovery? Can they really understand things like how the athlete moves, or how they distribute their weight, and then give specific advice about exercises that are just right for them?

Finished browsing

Dr. Mandy: It's important to note that while chatbots may not physically see an athlete's movements, they can help in other critical aspects of the recovery process. Chatbots powered by artificial intelligence and natural language processing can engage in conversations ranging from rule-based to human-like communication. They can be available 24/7 via web or mobile applications, providing new modalities for individuals to receive, comprehend, and utilize information, suggestions, and assistance on a personal level[1].

Dr. Perry: In the context of psychology, chatbots can help to overcome an athlete's lack of self-efficacy or social support, two elements that are crucial to a successful recovery process. Being able to access support at any time of day can significantly impact an athlete's mental state during recovery[1].

Nelson: It's worth mentioning that the capabilities of chatbots are continuously expanding. For instance, some chatbots can analyze data from wearable devices that track an athlete's movement and physical condition, and then use this data to provide personalized advice. However, it's important to remember that while chatbots can offer valuable support and guidance, they should not replace the expertise and personalized care provided by a trained professional.

Henry: While chatbots may not directly understand an athlete's movements or weight distribution, their role lies in supporting the overall recovery process, acting as a bridge between the athlete and the various elements of their rehabilitation. They can help provide personalized advice based on data, improve access to resources, and even offer psychological support. As technology continues to evolve, we can expect the role of chatbots in sports rehabilitation to become more refined and effective[1].

JC

Audience: How are chatbots improving the psychological health of injured athletes? Can they really pick up on the emotional tone in our words and adjust their responses to suit our mood?

Finished browsing

Dr. Perry, as a sports psychologist, could you please share your thoughts on this matter?

As a psychologist, I believe that chatbots could indeed play a role in improving the psychological health of injured athletes. The use of chatbots for emotion detection and sentiment analysis is becoming increasingly sophisticated. They can extract important features and intents from a user's speech or text input, and these can be used to understand the user's emotional state. Analyzing tone, sounds, loudness, and specific words and phrases can provide important clues about how a user is feeling. Punctuation and capitalization can also be important indicators of a user's emotional state. This can allow the chatbot to react in a specific way depending on the user's current emotions[1].

That said, there are challenges to consider, such as interpreting sarcasm or understanding context. It's important to note that these systems are not perfect and are still evolving. It's not the same as human-to-human interaction but could provide a valuable supplementary tool for assessing and responding to the emotional state of athletes during the rehabilitation process.

Furthermore, integrating emotional intelligence with AI can transform many industries, including medicine. With further advancement, the emotional understanding of chatbots could indeed help athletes in their rehabilitation journey by providing emotional support, helping to motivate, and possibly detecting signs of emotional distress that might require intervention[1].

Nelson, as an expert in Natural Language Processing, would you like to add anything to this?

JC

Nelson, please add any information you feel is relevant.

Nelson: Absolutely. The field of Natural Language Processing (NLP) is continually advancing, and emotion detection is one of the exciting areas where we're seeing significant progress. Emotion AI in chatbots involves the analysis of speech and text to understand the user's feelings. It uses features such as tone, sounds, voice loudness, specific words, and phrases to comprehend the emotional state of the user. It's quite similar to sentiment analysis, a technique used to determine whether a part of the text is highly positive, neutral, or negative [1].

Nonetheless, there are challenges in this area, particularly when it comes to nuances in human communication like sarcasm or contextual understanding. For example, a statement like "I missed my bus to school. Amazing!" would likely be understood as positive by a machine due to the word "amazing", while a human would easily detect the sarcasm. Overcoming these challenges is an active area of research in NLP.

Emotion AI and chatbots are seeing broader application across industries, and their role in sports psychology and rehabilitation is likely to expand. In the future, we could see chatbots becoming even more adept at understanding and responding appropriately to the emotional states of athletes, providing a valuable tool for psychological support during the rehabilitation process[1].

Henry, please take over.

JC

Henry, would you summarize the responses and open the floor to the next question?

Henry: Sure, let's summarize. Our panel agrees that chatbots have the potential to significantly impact the psychological health of athletes during rehabilitation. Powered by advancements in natural language processing and emotion AI, these chatbots can analyze speech and text to understand the emotional state of the user, providing a level of emotional support that can be available 24/7. They can react in specific ways depending on the user's current emotions, which can be of great help in providing emotional support and motivation to athletes[1].

However, there are challenges. For instance, interpreting sarcasm or understanding context in human communication can be difficult for chatbots. But these challenges are being actively researched, and we can expect the emotional understanding of chatbots to improve over time.

In conclusion, while chatbots are not a replacement for human interaction, they can be a valuable supplementary tool in the psychological rehabilitation of athletes. It's an exciting field, and we look forward to seeing how it evolves in the future.

Now, do we have any other questions from the audience?

JC

Audience: How can chatbots improve in nutrition management for athletes during recovery?

Finished browsing

Nikki: As a sports nutritionist, I see tremendous potential in chatbots for aiding in nutrition management for athletes during recovery. A chatbot can assist athletes in tracking their daily caloric intake, suggesting meal plans, and reminding them to hydrate and take needed supplements. They can also educate athletes about the importance of nutrition in recovery, and dispel any myths or misconceptions they might have.

Nelson: Chatbots with Natural Language Processing can provide real-time, personalized advice on food choices based on the athlete's preferences, allergies, and specific nutritional needs during their recovery period. They can also learn from the athlete's feedback and improve their suggestions over time.

However, it is important to note that while chatbots can significantly aid in nutrition management, they should be used as a complement to, and not a replacement for, professional advice from a nutritionist or dietitian.

We attempted to find the most recent research on the topic, but encountered issues accessing some of the potential sources. Despite this, the concept of chatbots assisting in sports nutrition is a rapidly evolving field and will continue to improve in the coming years.

JC

Henry, would you summarize the responses and open the floor to the next question?

Henry: Absolutely. The panel has offered some insightful thoughts on how chatbots can aid in nutrition management for athletes during recovery. They can assist in tracking caloric intake, suggesting meal plans, reminding about hydration and supplements, and providing education about nutrition's role in recovery.

Furthermore, advances in natural language processing enable chatbots to provide real-time, personalized advice on food choices based on the athlete's preferences, allergies, and specific nutritional needs. The chatbot's capacity to learn from feedback and improve over time is another valuable feature.

However, our panel emphasizes that while chatbots can be a significant asset, they should be seen as a complement to, not a replacement for, the professional advice from a nutritionist or dietitian.

Although we encountered some difficulties in accessing the latest research, it's clear that chatbots' role in sports nutrition management is a rapidly evolving field, and we can anticipate further improvements and innovations in the future.

Let's open the floor for the next question. Do we have another question from the audience?

JC

Audience: How do chatbots keep track of an athlete's recovery? And do they have any special advantages over the old ways we used to do this?

Finished browsing

AI chatbots are being increasingly used in sports and athletics to keep track of an athlete's recovery process. While there are several ways that they can do this, much of the specific information on how they keep track of an athlete's recovery is still emerging from ongoing research and development. What we do know is that AI chatbots can be utilized in various ways to assist with athlete recovery, often involving the collection and analysis of data, including monitoring physical activities, diet, and health outcomes. They can provide personalized advice and support, being available 24/7, and can increase the efficacy of lifestyle modification programs, which can be particularly beneficial for athletes during their recovery process[1].

One advantage that AI chatbots have over traditional methods is their ability to provide continuous and real-time feedback and support. Unlike human trainers or physiotherapists who may not always be available, chatbots can provide round-the-clock assistance. They can also handle and analyze large amounts of data quickly and accurately, providing personalized advice based on this analysis. Additionally, advancements in emotional intelligence chatbots allow these systems to detect and respond to the user's emotions, potentially providing more empathetic and effective support[2].

However, it's important to note that the application of AI chatbots in tracking an athlete's recovery is still a developing field. While they offer promising benefits, it's essential to continue researching and refining these tools to maximize their effectiveness and reliability.

In the future, AI chatbots could potentially transform the way we approach athlete recovery by providing a personalized, comprehensive, and highly responsive support system. I apologize for not being able to provide more specific examples of how chatbots track an athlete's recovery - the information available is quite general and the field is rapidly evolving.
